# Age-enhancing cognitive ability shows similar attenuation in task evoked brain networks with aging and preclinical AD

**DOI:** 10.1101/2025.11.02.25339341

**Authors:** Peter Chernek, Bardiya Ghaderi Yazdi, Seyed Hani Hojjati, Sindy Ozoria, Jenseric Calimag, Xiuyuan Hugh Wang, Ava Modarresi, Saman Gholipour Picha, Gloria Chiang, Qolamreza R. Razlighi

**Affiliations:** Quantitative Neuroimaging Laboratory of Brain Health Imaging Institute, Department of Radiology, Weill Cornell Medicine, New York City, United States of America; Brain Health Imaging Institute, Department of Radiology, Weill Cornell Medicine, New York City, United States of America; Fralin Biomedical Research Institute, Virginia Tech; Roanoke, United States of America

**Author notes:** These authors contributed equally to this work and share first authorship (Peter Chernek, Bardiya Ghaderi Yazdi).

**Keywords:** crystallized memory, fluid reasoning, resting-state functional connectivity, age-related, Aβ-related, BOLD Response

## Abstract

Brain aging - with and without pre-clinical Alzheimer’s disease (AD) pathology - are associated with deterioration in the brain networks’ coherence and/or co-activation/deactivation as well as with decline in most cognitive abilities, paving the road for a network-based conceptualization of the brain normal versus pathological aging. However, certain cognitive abilities, like crystallized memory, improve with age, which complicates the explanation of these changes solely through age-related decline in the brain networks. Using a cross-sectional cohort of 259 participants (62 young, and 197 older), which underwent two task-based (one declining and another improving with age), and one resting-state fMRI scans, plus a positron emission tomography scan (to determine preclinical amyloid accumulation), we found that the brain networks’ co-activation/deactivation, but not coherence, significantly attenuate with age and/or AD pathology even in the task for which performance improves by age. Interestingly, we also found that an increase in the networks’ co-activation/deactivation, but not coherence, was associated with an improvement in task performance. Finally, we provided preliminary evidence that the brain networks lose their task-evoked deactivations with age before their coherence. These findings shed light on the process of functional aging in the brain networks, differentiate functional aging of the brain networks’ coherence at rest versus their task-evoked co-activation/deactivation, and emphasize the more dominant role of the task-evoked brain activity in understanding aging brain function and distinguishing it from preclinical AD.

## Introduction

The human brain performs daily tasks through the coordinated inter-regional interaction of large-scale brain networks.^1^ Functional magnetic resonance imaging (fMRI) has been used to show that the regions of these networks co-activate/deactivate during various tasks performance, often referred to as task-evoked blood oxygenation level dependent (BOLD) response, and tend to work together in the absence of any external task, often referred to as resting-state functional connectivity (FC) networks.^1,2^ Both fMRI measurements have been shown to be associated with different cognitive abilities in healthy and clinical cohorts; opening the opportunity for network-based conceptualization of aging and Alzheimer’s disease (AD) which has recently gained tremendous attention in the field. It has been hypothesized that AD is a brain network disease that primarily deteriorates brain connections using different mechanisms.^3,4^ On the other hand, often decades before AD symptoms, about 25% of cognitively unimpaired (CU) healthy elderly individuals (>50 years) begin to show early deposition of AD pathology, Amyloid β (Aβ), which is now detectable *in-vivo* via positron emission tomography (PET) imaging.^5^ This offers a critical window for investigating the alterations and malfunctioning of brain networks and their relationship with future progress and symptoms of the disease.

The advent of fMRI revolutionized our understanding of the human brain function and discovery of various brain networks.^6^ While both task-evoked and resting-state functional networks have been instrumental in the understanding of the brain functional architecture, the relationship between these two fMRI measurements is still under investigation.^7,8^ Furthermore, both resting-state FC networks as well as task-evoked BOLD response particularly in the Default mode network (DMN) regions are shown to be disrupted in normal aging, asymptomatic/preclinical phase of AD, mild cognitive impairment (MCI), and AD.^9–12^ Despite this progress, the functional consequence of normal aging and preclinical AD across all the brain’s large-scale FC networks as well as their corresponding networks of task-evoked brain co-activation/deactivation is still under investigation.^7,13^

The DMN is one of the well-documented brain networks that has been studied with both task-evoked BOLD response as well as FC analysis.^1^ Anatomically, DMN comprises regions such as the posterior-cingulate/precuneus (PCC), inferior parietal, medial prefrontal cortex (mPFC), middle temporal, and hippocampus which are often active during rest and their activation decreases during almost any task.^14^ The decrease in the activation can be detected by task-evoked BOLD response in the opposite direction, which we term the “negative BOLD response” (NBR). ^7^ DMN regions have also shown strong FC during rest as well as during task-performance.^7,15^ Importantly, early Aβ deposition spatially overlaps with the DMN, which makes it the primary focus in many AD studies.^16^ Prior work from our group have already shown that these two overlapping fMRI measurements from DMN regions are dissociable and probably represent distinct underlying neurophysiological processes.^7^ However, age-related and AD-related alterations in the DMN task-evoked NBR and FC or in the other brain networks are yet to be fully or simultaneously examined.

Existing studies examining the functional alterations in the brain’s large-scale networks due to aging and preclinical AD often use resting-state FC networks, based on an underlying assumption that age and/or AD deteriorate brain networks’ functionality and subsequently lead to cognitive decline.^17–20^ However, the cognitive domains that show age-related improvement in performance pose a challenge for this hypothetical framework and highlight the need for investigating the brain networks’ task-evoked BOLD response, tapping into different cognitive domains particularly the ones showing age-related improvement.^21,22^ In this study, we propose to study brain networks FC at rest as well as the networks’ task-evoked BOLD response during two different tasks that have previously shown both age-related decline and improvement in performance. In addition, we propose to investigate all major brain networks together to observe a broader picture of the whole brain during aging and preclinical AD without limitation to the DMN alone. Our previous finding suggests that the FC networks provide the underlying functional architecture for the task-evoked networks which makes the FC networks indirectly involved in task performance. This aligns with the existing understanding that FC networks are representative of the brain’s on-going or spontaneous activities during rest, task, sleep, and even anesthesia.^23,24^ Therefore, we hypothesize that FC networks are extremely robust and resilient during aging and even during the preclinical stage of AD. Inversely, the task-evoked brain activation/deactivation networks, and in particular NBR within the DMN, will be disrupted by normal aging and will attenuate even further in preclinical AD groups. Consequently, we hypothesize that the subject-wise disruption in the brain networks’ task-evoked BOLD responses will precede the alteration in the same networks FC suggesting it as a more reliable and earlier biomarker of the aging and preclinical AD. Finally, based on our previous findings, we hypothesis that the increase in the brain networks BOLD responses, unlike their FC, will be associated with better task-performance in both cognitive domains.

## Results

### Cohort demographic and characteristics

The demographics and characteristics of our study participants are listed in Table 1. The study recruited 261 participants, categorized into two groups based on age. Fourteen participants were excluded for not having amyloid FBB PET, reducing our total sample size to 247 participants. The younger group comprised 62 individuals aged 20-40 years (mean ± SD = 27.8 ± 5.3). The elderly group included 185 individuals aged 60-80 years (mean ± SD = 68.5 ± 5.7). We further separated elderly group into HE (Aβ –; *N*=141; age=67.6 ±5.5 years) where no Aβ deposition was detected by visual reading of the FBB PET scans and CU (Aβ +; *N*=44; age=71.5 ±5.5 years) group where the presence of Aβ in FBB PET scan was confirmed by visual reading. The incident rate of Aβ positivity in our elderly cohort was 24% which is comparable to existing reports. CU elderly group was on average older than HE and the difference was significant (Δage=3.8 years, p=0.0001). Since some of the younger participants were still in school, the average education in the young group (16.5 ± 2.0 years) was slightly lower than both elderly groups (HE: 17.0 ± 2.2 years; CU: 17.1 ±2.7 years), but neither difference was significant (HE vs. HY: 0.5 years, p=0.57 CU vs HY: 0.6 years, p=0.58), nor was the educational difference between HE, and CU (0.1 years, p=1.0). In the total study population, 49% (*N*=121) were female and there were no significant differences in the female percentage between the three groups (HY: 48%; HE: 48%; CU:52%). There were significant differences in the ethnic makeup between both elderly groups and HY. HY showed more diverse ethnic makeup (53.2% white) than elderly groups did (HE: 85.1% white, CU: 86.4 % white). As expected, both CU and HE showed significantly reduced MoCA scores compared to HY (CU: (18.9 ± 2.0), HE: (19.7 ± 1.8)) versus (HY: 20.9 ± 1.1). However, MoCA scores between CU and HE groups were not different after adjusting for age and multiple comparison correction (ΔMoCA= -0.9, *p*=0.117). Mean Global Aβ SUVR increased by group from HY (1.06 ±0.02) to HE (1.09± 0.06) to CU (1.49 ±0.32). As expected, only CU elderly group showed a significantly higher Global Aβ in comparison to HY, and HE (CU vs HY: ΔAβ=0.43, *p*<0.0001; CU vs HE: ΔAβ=0.40, *p*<0.0001) and the difference between two elderly group was no significant (HE vs HY: ΔAβ=0.03, *p*=0.49). Finally, in agreement with the existing reports, the performance in the antonyms task improved significantly by age for both elderly groups compared to HY (HE: ΔAcc/RT =0.03, *p*=0.0004; CU: ΔAcc/RT =0.03, *p*=0.024) whereas the performance on the matrix reasoning task significantly deteriorated by age for both elderly groups compared to HY (HE: ΔAcc/RT = -0.02, *p*=0.000; CU: ΔAcc/RT= -0.02, *p*=0.000). There were no significant Aβ-related differences found between the performance of the two elderly groups (HE and CU) for either antonyms or matrix reasoning tasks.

**Table 1.** Demographic and Characteristics of Study Cohort Stratified by age and Amyloid PET findings.

| **Total N**<br>**(247)** | **HY**<br>**62** | **HE (Aβ -)**<br>**141** | **CU (Aβ +)**<br>**44** | **CU vs. HY**<br>**Mean Diff (p)** | **HE vs. HY**<br>**Mean Diff (p)** | **CU vs. HE**<br>**Mean Diff (p)** |
| --- | --- | --- | --- | --- | --- | --- |
| **Age**<br>(Mean±SD<br>yrs)<sup>a</sup> | 27.8 ±5.3 | 67.6 ±5.5 | 71.5 ±5.5 | 43.6 (0.000) | 39.7 (0.000) | 3.8 (0.0001) |
| **Gender**<br>Female (%)<br><sup>b</sup> | 30(48%) | 68 (48%) | 23(52%) | 4% (1.000) | 1% (1.000) | 4% (1.000) |
| **Education**<br>**(yrs) <sup>b</sup>**<br>Mean ±SD | 16.5 ±2.0 | 17.0 ±2.2 | 17.1 ±2.7 | 0.6 (0.580) | 0.5 (0.570) | 0.1 (1.000) |
| **Ethnicity <sup>b</sup>**<br>Count(%) |  |  |  |  |  |  |
| Caucasian | 33(53.2%) | 120(85.1%) | 38(86.4%) | 33.2% (0.005) | 31.9% (0.000) | 1.3 (1.000) |
| Hispanic | 9(14.5%) | 1(0.7%) | 0(0.0%) |  |  |  |
| AA | 7(11.3%) | 10(7.1%) | 4(9.1%) |  |  |  |
| Asian | 9(14.5%) | 6(4.3%) | 0(0.0%) |  |  |  |
| Other | 4(6.5%) | 4(2.8%) | 2(4.5%) |  |  |  |
| **MoCA<sup>b</sup>**<br>Mean ±SD | 20.9 ±1.1 | 19.7 ±1.8 | 18.8 ±2.0 | -2.1 (0.000) | -1.2 (0.000) | -0.9 (0.117)<sup>c</sup> |
| **Global Aβ<sup>b</sup>**<br>(SUVR<br>Mean ±SD) | 1.06 ±0.02 | 1.09± 0.06 | 1.49 ±0.32 | 0.43 (0.000) | 0.03 (0.490) | 0.40 (0.000) |
| **Antonyms**<br>Acc/RT<br>(Mean<br>±SD) | 0.098±<br>0.037 | 0.126 ±<br>0.048 | 0.123 ±<br>0.046 | 0.025 (0.024) | 0.028 (0.0004) | -0.003 (1.000) |
| **Matrix**<br>**Reasoning**<br>Acc/RT<br>(Mean<br>±SD) | 0.039±<br>0.013 | 0.023 ±<br>0.009 | 0.022 ±<br>0.009 | -0.017 (0.000) | -0.016 (0.000) | -0.001 (1.000) |

Abbreviations - HE: Healthy Elderly, HY: Healthy Young, CU: Cognitively Unimpaired, Aβ +: Amyloid Positive, Aβ -: Amyloid Negative, Acc: Accuracy, RT: Response time, AA: African American, <sup>a</sup>Welch 2 sample t test, <sup>b</sup>Chi-square test, <sup>c</sup>Adjusted for age,
<sup>1</sup>Comparison of Aβ + CU to HY group, <sup>2</sup>Comparison of Aβ – HE to HY group, <sup>3</sup> Comparison of Aβ + CU to Aβ – HE group, All pairwise group comparisons adjusted for multiple comparisons using Bonferroni correction method.

### Alteration in BOLD response across aging and preclinical AD

In this experiment we have shown that task-evoked BOLD response significantly changes during normal aging and in preclinical AD, whereas FC remain almost intact. We selected two fMRI-compatible tasks from two cognitive domains (crystalized memory and fluid reasoning) which have opposite age effects in performance.^25^ As shown in Table 1, while elderly’s performance improves by age during antonyms, it decreases during matrix-reasoning. Figure 1a illustrates the vertex-wise magnitude of the mean BOLD response color-coded and projected on the surface of the semi-inflated cortex in MNI space for HY, HE, and CU groups (ordered from left to right) during antonyms and Figure 2a shows the same for matrix-reasoning. As illustrated, all three groups demonstrate a significant NBR pattern covering the DMN regions and a significant PBR pattern covering the DAN and other task-positive networks. In addition, a strong PBR in the visual cortex due to visual representation of the task as well as a strong PBR in the left motor cortex due to participants’ response with their right hand is noteworthy and anticipated for all three groups. The attenuation in the magnitude of the NBR from HY to HE and even from HE to CU groups can be seen whereas no difference in the PBR between the three groups can be identified visually. Vertex-wise linear regression analysis was used to examine the statistical difference between the mean of the BOLD response magnitude between the groups while controlling for age when comparing HE and CU. Figures 1b/2b show the results of this experiment by projecting the vertex-wise t-stats of the significant difference on the surface of semi-inflated cortex for antonyms/matrix-reasoning task when comparing HY vs HE, HY vs CU, and HE vs CU, respectively from left to right. As expected, both tasks demonstrate a significant age-related and Aβ-related decrease in the magnitude of the NBR in almost all DMN regions.

**Figure 1.**
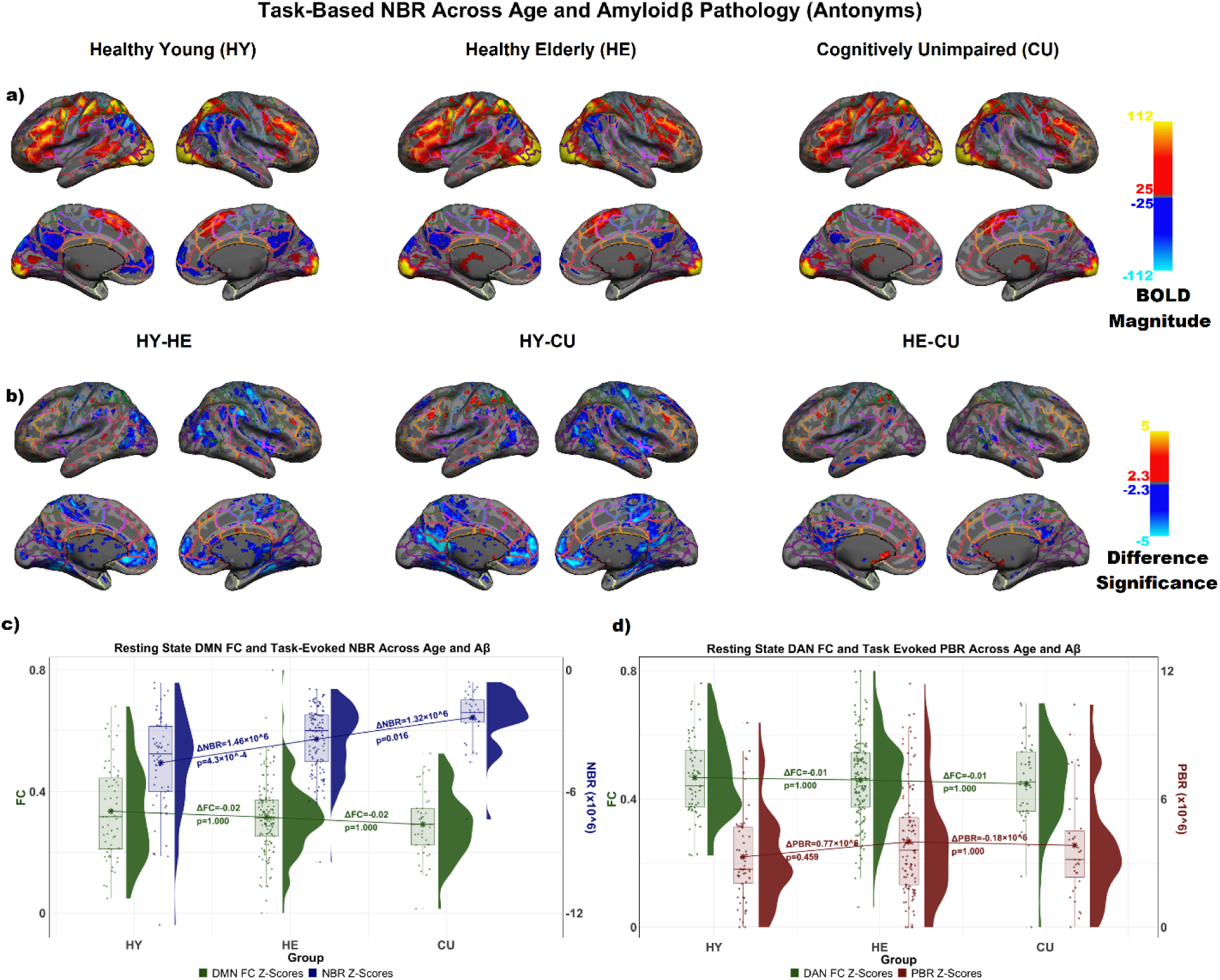
Task-evoked negative BOLD response deteriorates with age and preclinical Aβ deposition during antonyms task. **a)** depicts the magnitude of the group-wise mean BOLD response color-coded (red∼yellow for PBR and blue∼light-blue for NBR) and projected on the surface of a semi-inflated cortex in MNI space for HY, HE, and CU groups respectively from left to right. The borderlines delineate the Schaefer atlas regions and borderlines’ color indicate the Schaefer atlas networks (DMN: red, DAN: orange); **b)** depicts the t-stats of the vertex-wise multiple linear regression for comparing the pair-wise group mean differences in the BOLD response magnitude which are color-coded and projected on the surface of semi-inflated cortex in MNI space for HY-HE , HY-CU, and HE-CU, respectfully from left to right; **c)** Rain-cloud plots depict the distribution of the subject-wise negative BOLD response (in blue) and FC (in green) z-scores within the DMN for HY, HE, and CU groups. Between group group differences are shown on connecting lines; **d)** Rain-cloud plots depict the distribution of the subject-wise positive BOLD response (in red) and FC (in green) z-scores for DAN in HY, HE, and CU groups. Between group differences are shown on connecting lines;

**Figure 2.**
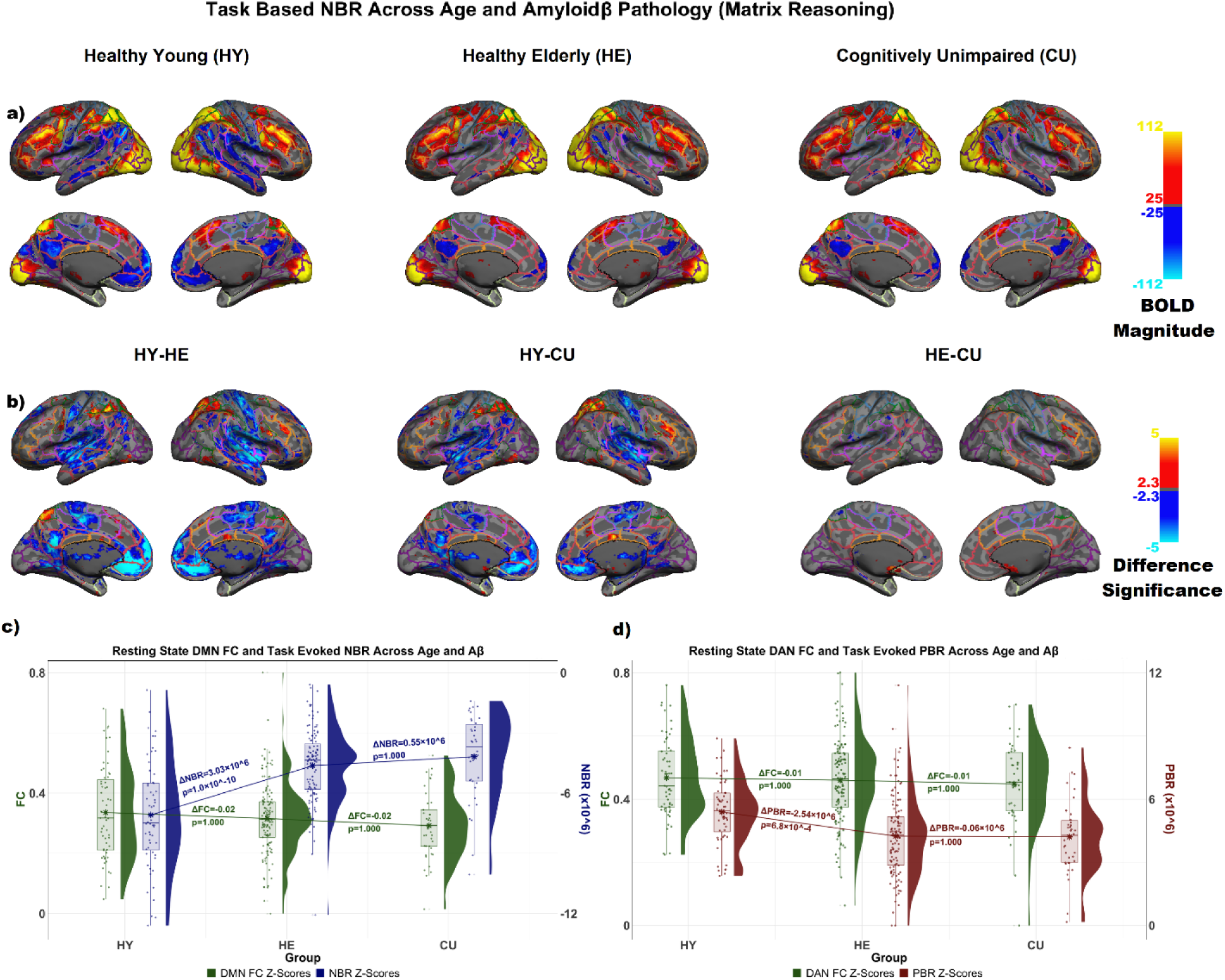
Task-evoked BOLD response decreases substantially with age, but not with preclinical Aβ deposition during matrix-reasoning task; **a)** depicts the magnitude of the group-wise mean BOLD response color-coded (red∼yellow for PBR and blue∼light-blue for NBR) and projected on the surface of a semi-inflated cortex in MNI space for HY, HE, and CU groups respectively from left to right. The borderlines delineate the Schaefer atlas regions and borderlines’ color indicated the Schaefer atlas networks (DMN: red, DAN: orange); **b)** depicts the t stats of the vertex-wise multiple linear regression for comparing the pair-wise group mean differences in the BOLD response magnitude are color-coded and projected on the surface of semi-inflated cortex in MNI space for HY-HE , HY-CU, and HE-CU, respectfully from left to right; **c)** Rain-cloud plots depict the distribution of the subject-wise negative BOLD response (in blue) and FC (in green) z-scores for DMN in HY, HE, and CU groups. Between group differences are shown on connecting lines; d) Rain-cloud plots depict the distribution of the subject-wise positive BOLD response (in red) and FC (in green) z-scores for DAN in the HY, HE, and CU groups. Between group differences are shown on connecting lines.

Notably, the age-related reduction in the NBR is more prominent in the posterior cingulate and medial orbito-frontal regions of the DMN for both antonyms and matrix-reasoning. However, the difference in the NBR of the DMN was more significant for matrix reasoning task. There is only a small and negligible decrease in the PBR of the DAN due to age or Aβ pathology for antonyms, which became more prominent during matrix-reasoning task. Age and Aβ-related increase in the magnitude of the NBR and PBR is almost non-existent.

### Quantify the age-related and A**β**-related alterations in the task-evoked BOLD response

Within each brain network, we summed the magnitude of all the voxels that showed a significant positive or negative BOLD response (|t|>5) during each task separately. This method quantifies both expression and extent of the BOLD response in each network. To compare alteration in the task-evoked BOLD response with the alterations of the same network’s resting-state FC, we plotted the three groups’ subject-wise distribution of the BOLD response and FC side-by-side for DMN (Figures 1c/2c), and DAN (Figures 1d/2d). Each networks subject-wise FC were computed by averaging all within network pair-wise correlations. Figures 1c/2c depict the distribution of the subject-wise NBR obtained from the DMN regions separately for each group (HY, HE, and CU, respectively from left to right) using a blue-colored rain-cloud plot. For comparison, the distribution of the subject-wise FC from the same group and same network (DMN) is also plotted next to the NBR using a green-colored rain-cloud plot.

For antonyms task, participants with excessive head motion (scrubbed more than 30%) were excluded from this experiment (9 HE and 6 CU). One way ANOVA indicates that the NBR from DMN changes significantly with age and with Aβ (F=17.59, *p*<0.001). All three groupwise comparisons in NBR were statistically significant after Bonferroni multiple comparisons correction (HE vs. HY: ΔNBR=1.46x10^6^, *p*<0.001, CU vs. HY: ΔNBR=2.79x10^6^, *p*<0.000, CU vs. HE: ΔNBR=1.32x10^6^, *p*=0.01) (Figure 1d). On the other hand, one way ANOVA indicates that there is no significant difference in group means of the PBR from DAN (F= 1.6, *p*=0.205) (Figure 2d). For comparison, we also depict the distribution of the subject-wise FC for the DAN next to the distribution of the PBR (Figure 2d) reinforcing that there is no age-related or Aβ-related change in the FC within the DAN. We repeated these analyses by removing the outliers which did not change significance of the results. Similar results were obtained for Limbic (LIM), Ventral Attention Network (VAN), and Frontoparietal Network (FPN) and are depicted in Supplementary Figures 1, 2, and 3. These results provide strong evidence that normal aging and preclinical AD are associated with a reduction in the NBR with corresponding stability of the FC and PBR during antonyms task.

For matrix reasoning, participants with excessive head motion (scrubbed more than 30%) were also excluded (17 HE and 5 CU). One way ANOVA indicates that there is a significant difference in group means of the DMN NBR (F= 30.57, *p*<0.001) and DAN PBR (F= 8.49, *p*<0.001). As shown in Figures 2c and 2d, age-related groupwise comparisons in DMN’s NBR and DAN’s PBR revealed a statistically significant age-related differences after Bonferroni multiple comparison correction (NBR: HY vs. HE: ΔNBR=3.03x10^6^, *p*<0.000, HY vs. CU: ΔNBR=3.58 x10^6^, *p*<0.000; PBR: HY vs. HE: ΔPBR=-2.54x10^6^, *p*<0.000, HY vs. CU: ΔPBR=- 2.6x10^6^, *p*<0.000); Healthy aging revealed a more prominent attenuation of the NBR for matrix reasoning when compared to antonyms. However, Aβ was not associated with alteration in NBR and PBR. As shown previously, the FC strength in the DMN and DAN remains stable from HY to HE to CU as depicted along with the distribution of the NBR and PBR for comparison in Figures 1d and 2d. We repeated these analyses by removing the outliers which did not change the significance of the results. Results obtained for LIM, VAN, and FPN are depicted in Supplementary Figures 4, 5, and 6 respectively. As depicted, FPN also shows a significant reduction in PBR for CU compared to HY, whereas LIM shows a significant increase in PBR from HY to HE.

Altogether, these results suggest that age-related change in the performance of the tasks is somewhat independent of the age-related changes in the brain networks task-evoked BOLD response and FC. This is because, while the age-related change in performance of the two tasks is in opposite directions, the age-related change in the task-evoked BOLD response were in the same direction and no change has been identified for the networks’ FC. In addition, age-related and Aβ-related alterations in the functionality of the brain network are more evident and detectable in the task-evoked BOLD response than in their FC.

### Robustness of FC networks across aging and preclinical AD

In this experiment, we provide evidence that FC networks are robust throughout the aging, and they remain stable even in preclinical AD with confirmed AD pathology. To do this we computed pairwise inter-regional FC between the timeseries of 200 regions of Schaefer atlas for each participant, resulting in a 200x200 FC matrix. We compared the mean inter-regional connectivity between the three groups (HY, HE, and CU). Participants with excessive head motion (scrubbed more than 30%) were excluded from this experiment (1 HY, 2 HE, and 3 CU). Figure 3a illustrates the mean inter-regional FC (quantified with PCC for all possible regional pair timeseries) color-coded and depicted as a separate correlation matrix for HY, HE, and CU groups, respectively from left to right. As shown, all three groups represent higher FC within the 7 networks as they cluster together within each network along the main diagonal of the correlation matrix. Additionally, the two off-diagonal clusters of high FCs are the inter-hemispheric correlation within each network. The anti-correlated networks are also clearly shown in the figure with negative PCCs which are color-coded in blue. Qualitatively, there are no visually detectable differences between these three FC matrices. Supplementary Figure 7 depicts the results after Bonferroni multiple comparisons correction of the statistical testing for significant differences in the mean of the inter-regional FC between the three groups controlling for motion (FD) (from left to right: HY-HE, HY-CU, and HE-CU). While there are some significant differences in the inter-network FC, only four differences in the within networks FC can be observed; one reduction in the DMN inter-hemispheric pair (HY vs. HE; LH_Default_Temp_4 & RH_Default_Temp_3; ΔFC=0.27, p=0.026), one reduction in FPN within hemispheric pair (HY vs. CU; LH_Cont_Par_2 & LH_Cont_PFCl_5; ΔFC=0.231, p=0.03), one reduction in the Limbic within hemispheric pair (HY vs. HE; RH_Limbic_TempPole_1 & RH_Limbic_TempPole_2; ΔFC=0.246, p=0.039), and one reduction in Limbic inter-hemispheric pair (HY vs. HE; LH_Limbic_TempPole_2 & RH_Limbic_TempPole_1; ΔFC=0.222, p=0.017). Most of these regions are in the temporal lobe and more toward medial side and temporal pole which has shown a high level of fMRI air-susceptibility artifact and age-related brain atrophy, highlighting the possibility of the fMRI signal drop-out in elderly group.^26^ In addition, all four significant findings disappeared when we repeated this analysis with participants having less motion (scrubbed less than 10%, 5%, and 1%), highlighting the possibility of motion driving the effect and re-iterating the substantial effect of motion on FC studies.^27,28^

**Figure 3.**
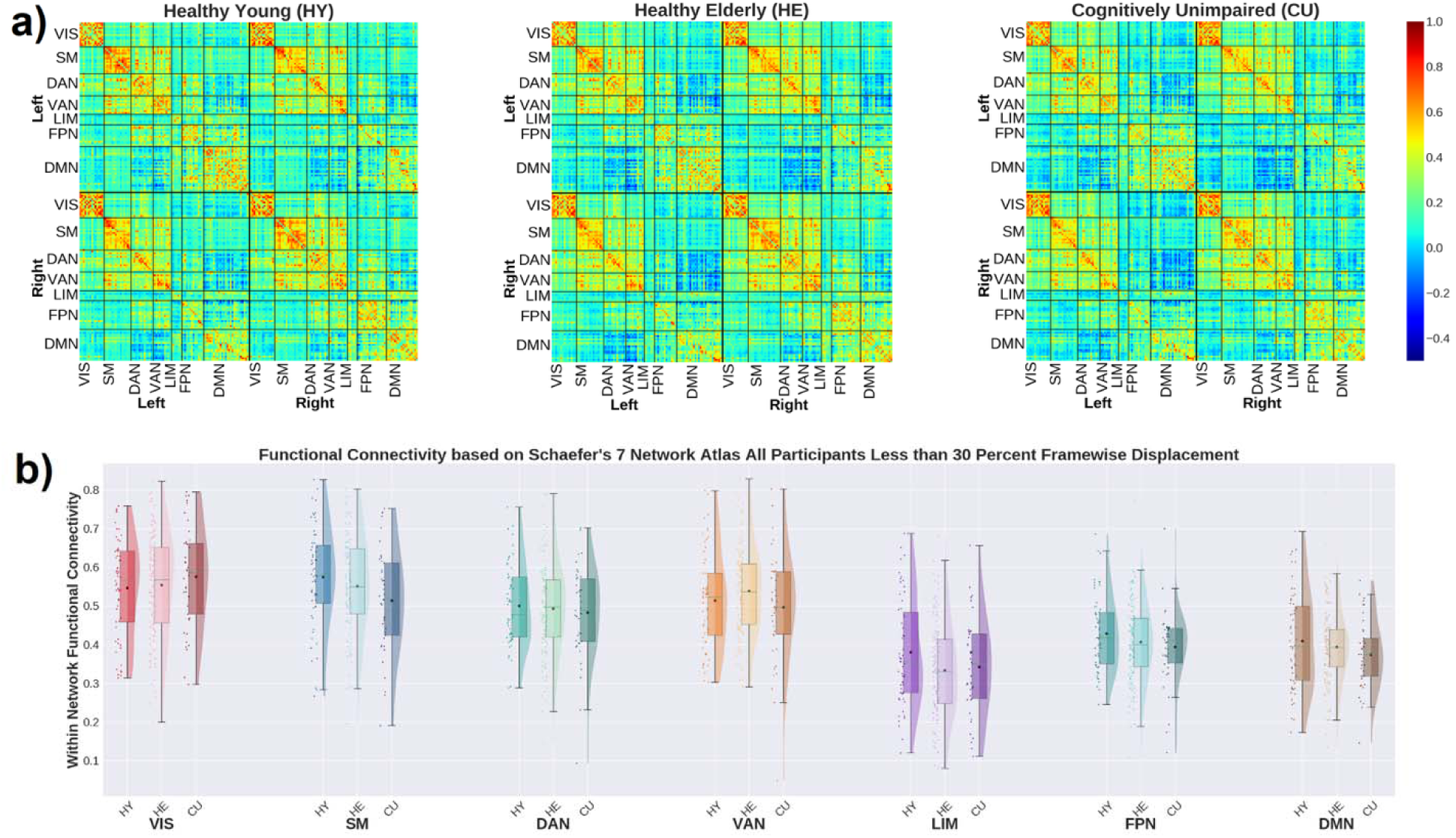
Robustness of functional connectivity network during aging and preclinical AD; **a)** Mean inter-regional FC of the 200 Schaefer regions resulting in 200 x 200 correlations maps organized by the seven major brain networks color-coded and plotted from left panel for HY, middle panel for HE, and right panel for CU**, b)** Illustrates the comparison between the distribution of the subject-wise FC across the three groups (HY, HE, And CU) separately for each of 7 brain network’s from Schaefer Atlas.

Next, we computed the within network connectivity by averaging all pairs of connectivity within each network, excluding interhemispheric pairs, and comparing them across the three groups (HY, HE, and CU). We showed the results of this experiment for the DMN and DAN alongside the task-evoked BOLD response in the previous section. In Figure 3b, we illustrate the results of this experiment for all seven of the major brain networks from the Schaefer atlas using 3-group rain cloud plots. Using ANCOVA and adjusting for FD and age (only when comparing HE to CU) we found no difference in within network FC between HY and HE, HY and CU, or HE and CU after Bonferroni multiple comparisons correction. We repeated this analysis by removing outliers which did not change the results. Repeating the analysis for participants with less head motion (scrubbed less than 10%, 5%, and 1%) resulted again in a similar finding (no age-related or Aβ-related change in the within network FC (see Supplementary Figures 8 and Supplementary Table 1 for details of this replication analysis within the DMN). These results show a strong pattern of stability of the 7 major brain networks across age and pre-clinical accumulation of Aβ pathology in terms of overall functional connectivity.

### Within-subject comparison of the FC and BOLD response

The previous experiment was mainly focused on the comparison of group differences where individualized measurements of within-subject comparisons were ignored. The goal of this experiment was to show that in comparison to HY, each elderly individual demonstrates more age-related and Aβ-related disruption in the NBR/PBR than in FC of the same network. To compare within-subject alteration in the task-evoked BOLD response directly with strength of the same network’s resting-state FC, we used young groups as our normative reference group to convert all participants BOLD responses and FC to z-scores. Essentially, the z-scores of the NBR/PBR and FC represent severity of age-related or Aβ-related alterations within functional networks in a scale that are comparable directly to each other. The distribution of the z-scores depicted using rain-cloud plots in Figures 4a/4c for DMN and 4b/4d for DAN for both antonyms and matrix-reasoning tasks. As shown in previous experience, while the FC strength remained stable across HY to HE to CU, the magnitude of the NBR decreased almost half a standard deviation from HY to HE and almost another half to CU for antonym task whereas for matrix reasoning the magnitude of the NBR decreased almost one standard deviation with age, but further attenuation by pre-clinical amyloid (Aβ) was not observed. The statistical significance for within subject differences were computed using a mixed-design repeated measure ANOVA to assess the within subject differences between disruptions of NBR versus FC in the DMN as well as disruption of PBR and FC in DAN. Controlling for age, there were highly significant differences between disruptions of NBR and FC z-scores in the DMN for antonyms (F=141, *p*<0.0001) as well as matrix-reasoning (F=352, *p*<0.0001), suggesting that the disruption of the NBR is significantly higher than FC in each participant. These differences were also interacting by Aβ positivity for antonyms (F=15, *p*<0.001) but not for matrix-reasoning (F=0.06, *p*=0.1).

**Figure 4.**
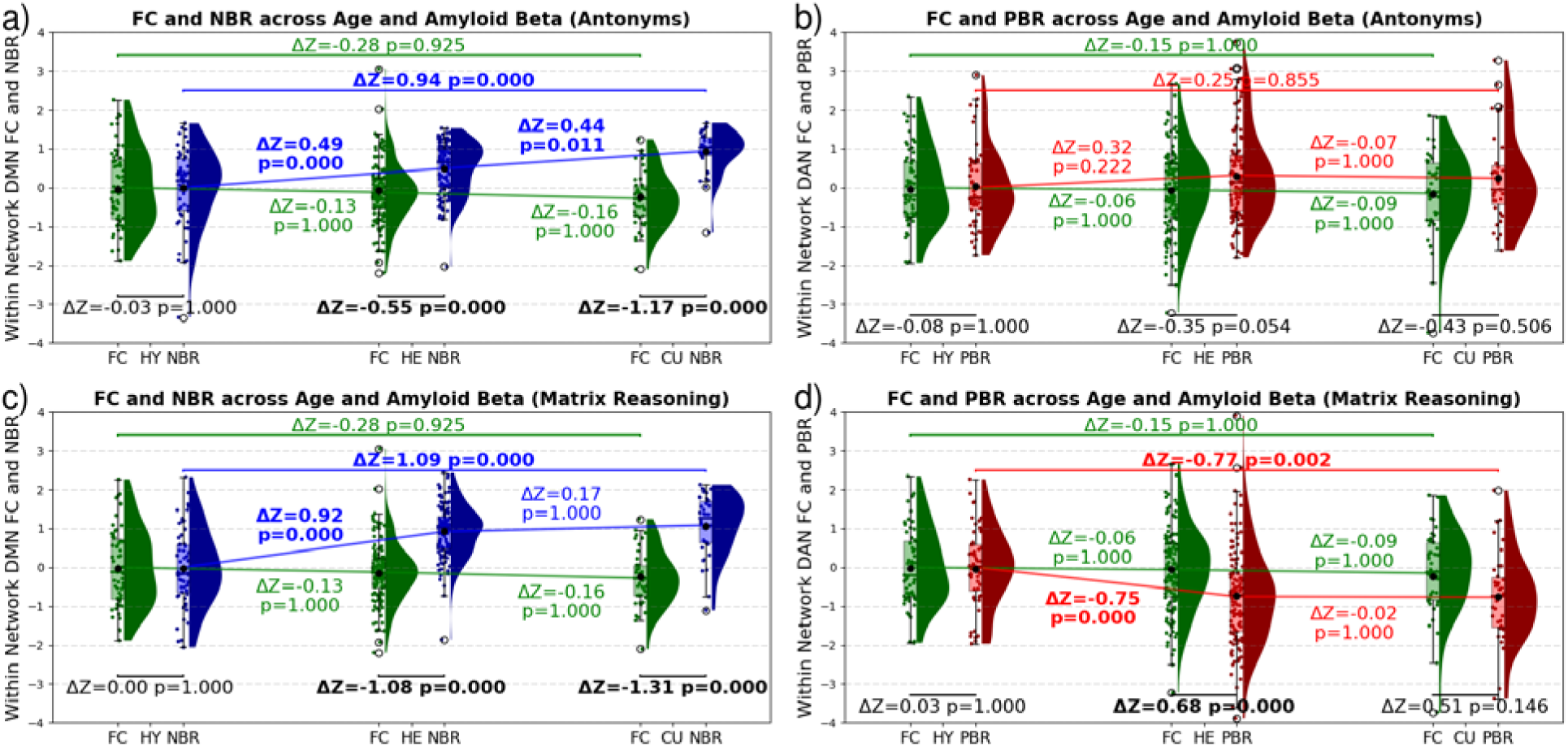
Comparisons of the within-subject alterations in the BOLD response and FC from DMN and DAN due to age and preclinical Aβ deposition; **a)** Rain-cloud plots depict the distribution of the subject-wise NBR (in blue) and FC (in green) z-scores for DMN in the HY, HE and CU groups in antonym task**. b)** Rain-cloud plots depict the distribution of the subject-wise PBR (in red) and FC (in green) z-scores for DAN in the HE and CU groups in antonym task. **c)** the same as **(a)** for matrix reasoning task, **d)** the same as **(b)** for matrix reasoning task. The statistical significance of the pairwise within-subject comparisons of the networks BOLD response and FC are given in black. The statistical significance of the between groups comparisons are given in the corresponding colors. We should emphasize that positive NBR z-scores correspond to disturbed NBR (lower magnitude of NBR) and negative NBR z-scores correspond to increased magnitude of NBR. The normative samples (HY) distributions of the z-scores are also depicted only for illustration of the reference points.

The within subject differences between the PBR and FC of the DAN were also significant for antonyms (F=15, *p*<0.01) and matrix-reasoning (F=49, *p*<0.0001) but the differences did not interact with Aβ for either task (F<1, *p*>0.1). As seen in Figures 4a/4c, the post hoc pair-wised t-tests for the group differences revealed a significant difference between DMN NBR and FC z-scores (antonyms: ΔZ=-0.55, *p*<0.0001, matrix-reasoning: ΔZ=-1.08, *p*<0.0001) which increased by presence of Aβ (antonyms: ΔZ=-1.17, *p*<0.001; matrix-reasoning: ΔZ=-1.31, *p*<0.001). In addition, as shown in Figure 4b, the post hoc pair-wise t-tests for group differences showed no significant difference between DAN PBR and FC z-scores for antonyms (ΔZ=-0.35, *p*>0.054) even in the presence of Aβ (ΔZ=-0.43, *p*>0.5). However, for matrix-reasoning, there was a significant difference between DAN PBR and FC z-scores (ΔZ=0.68, *p*<0.001), but in the presence of Aβ this difference was not seen. (ΔZ=0.51, p>0.1) (Figure 4d) Altogether, these results suggest that within the DMN, individual participants experience age-related disruption in their NBR, and the disruption worsens by presence of Aβ whereas FC remains relatively stable.

### Temporal ordering of the disruptions of the NBR and FC in the DMN

In this study, we also provide preliminary evidence of the temporal ordering of the events in normal aging and pre-clinical AD, even though our cohort was cross-sectional. We used two different analyses for this experiment. The scatter plots in Figures 5a/5b show the relationship between the subject-wise z-scores of the NBR and FC in the DMN during antonyms and matrix-reasoning tasks, respectively. Each point represents a single elderly participant. Again, as NBR values are negative, more positive NBR z-scores correspond to more disturbed NBR (less magnitude of NBR values) and more negative NBR z-scores correspond to increased magnitude of NBR. As shown, a greater proportion of participants had z-scores for NBR greater than z-scores for FC indicating a disruption in NBR prior to disruption in FC. A permutation test indicates that the number of participants with -z(NBR)<z(FC) is significantly higher than the number of participants with -z(NBR) >z(FC) (permutation test: antonyms: *p*<0.0001; matrix-reasoning: *p*<0.0001). Next, we used the bootstrap resampling of the posterior probabilities to show that the probability of observing disruption in NBR when FC is already disrupted is significantly higher than the probability of observing the disruption in FC when NBR is already disrupted. Figures 4c/4d show the results of this experiment for antonyms and matrix-reasoning tasks, respectively. As shown, the probability of observing disruption in NBR (lower magnitude of NBR) when FC is already disrupted (reduced FC) is significantly higher than the probability of observing disruption in FC when NBR is already disrupted (antonyms: *p*<0.001; matrix-reasoning: *p*<0.001). Altogether these results provide evidence that disruption in the NBR during aging and preclinical AD occurs prior to the disruption in the FC within the DMN.

**Figure 5.**
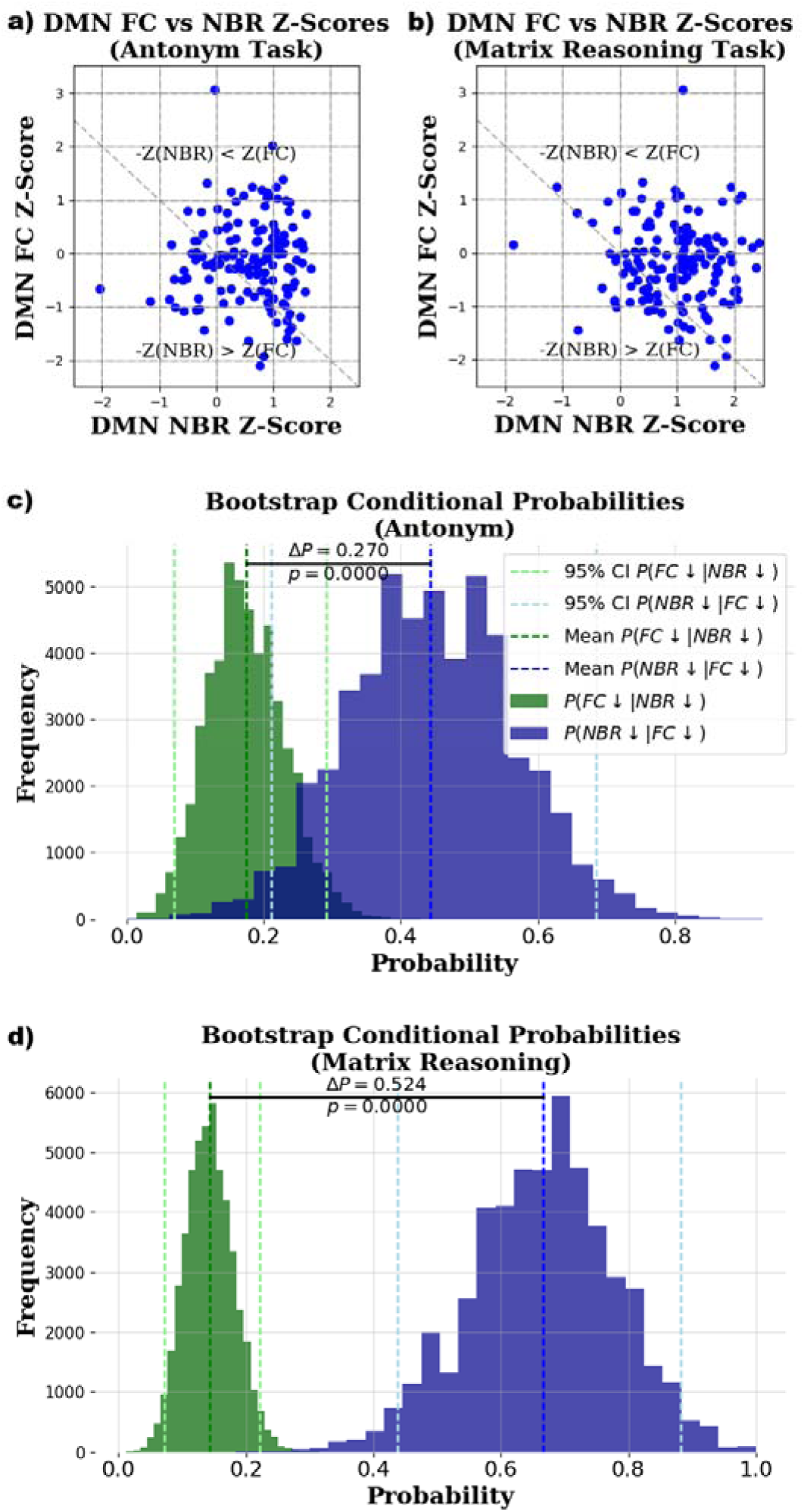
Age-related disruption in the task-evoked BOLD response of the DMN precedes alteration in its FC during both antonym and matrix-reasoning task; scatter plots depicting the relationship between age-related attenuation in the DMN’s NBR versus its FC for **a)** antonym and **b)** matrix reasoning tasks. Each points represent a single elderly participant. The histograms depict the results of the bootstrapping that showed the probability of observing disruption in NBR when FC is already disrupted is significantly higher than the probability of observing disruption in FC when NBR is already disrupted during **c)** antonym and **d)** matrix reasoning tasks.

### Association of BOLD response and FC with performance

The aim of this experiment was to show that task-evoked BOLD responses of the brain networks within elderly are more closely associated with task performance than underlying FC or global amyloid-Aβ during pre-clinical AD. As shown in Figures 6a/6b, after controlling for age and gender, an increase in the magnitude of DMN’s NBR (β=-4.96, *p*=0.01) and DAN’s PBR (β=4.5, *p*=0.001) was associated with better performance for antonyms task. However, neither mean DMN FC nor global Aβ deposition revealed a significant relationship with performance (Figures 6c/6d). Similarly, for matrix-reasoning, an increase in the magnitude of DAN’s PBR (β=4.38, *p*=0.016) correlated with better performance however this relationship was not observed with the DMN’s NBR. Again, neither mean DMN FC nor global Aβ deposition showed a significant relationship with performance for matrix-reasoning (Figures 5c/5d). Altogether, these results suggest that independent of the age effect on task performance, an increase in the magnitude of the BOLD response in the brain networks results in better performance whereas the same networks mean FC and global Aβ deposition do not correlate well with performance.

**Figure 6.**
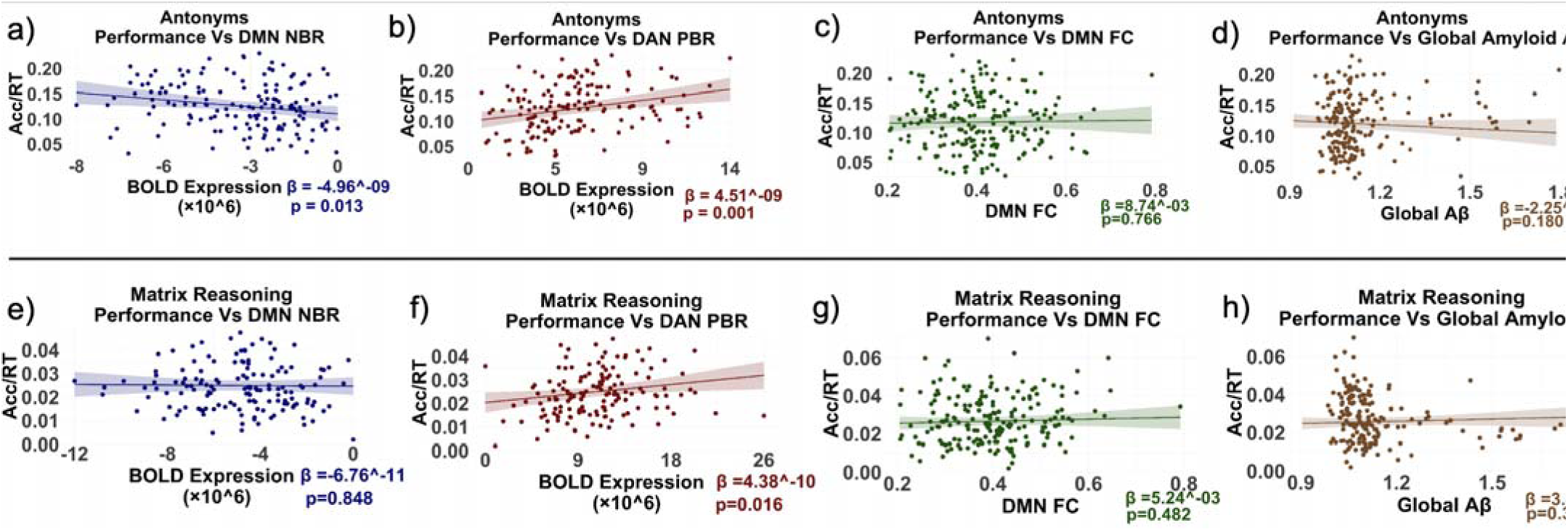
Task-evoked BOLD responses of the brain networks’, unlike their FC, are closely associated with task performance; Scatter-plots depict the association between the antonym task performance (measured by accuracy/RT) and **a)** NBR from the DMN regions, **b)** PBR from the DAN regions, **c)** FC of the DMN, and **d)** Global Aβ SUVR. Scatter-plots from **(e)** to **(h)** depict the same for matrix-reasoning task. The solid lines are the slope of the multiple regression analysis when age and gender are the co-variates.

## Discussion

Our results show that although the FC of the brain’s large-scale networks remains stable through aging and pre-clinical stage of AD with confirmed Aβ deposition, NBR from the DMN regions - during antonyms task - is attenuated by aging and even more with Aβ accumulation, revealing not only a close interplay between DMN’s NBR and Aβ independent of aging, but also NBR as an earlier brain biomarker for aging and preclinical AD. Interestingly, during the same task, PBR from a task-positive network, DAN, remained stable with aging and Aβ positivity. Other major task-positive brain networks (FPN, VAN, and LIM) showed similar results. Performing the same analysis for matrix-reasoning task, where performance declines with age, resulted in an even more prominent age-related attenuation in the NBR from DMN; However, the Aβ-related deterioration in the DMN’s NBR was not significant in comparison to CU anymore. Instead, for this difficult task, PBR from the DAN network showed significant age-related but not Aβ-related attenuation. Furthermore, we provide evidence that within-subject age-related attenuation in the task-evoked NBR not only is more prominent than alteration in FC but also seems to precede the alteration in the same network FC. Finally, the task evoked BOLD response should be investigated more closely in future studies on aging and pre-clinical AD due to the strong relationship observed with performance which was not seen with either FC or global Aβ.

### Alteration in BOLD response across aging and preclinical AD

The DMN’s NBR was significantly affected by both age and Aβ pathology for antonyms task which its performance improves by age. The magnitude of NBR decreased stepwise: decreases from HY to HE and further attenuates within CU. Notably, Aβ presence weakened the DMN’s NBR even after adjusting for age, suggesting a direct link between Aβ and reduced NBR in CU aligning with previous reports in both normal aging^9,12,20,29–39^ and pre-clinical AD.^12,30,39,40^ Even elderly without Aβ deposition revealed a reduced deactivation^30,31,38–40^ Higher NBR magnitude of DMN was associated with better performance, replicating prior findings.^37,39,41,42^ Some studies have shown a quadratic relationship between Aβ and NBR as participants with slightly elevated SUVR show reduced NBR magnitude while those with lower and higher SUVR show higher magnitude of DMN NBR.^35^ These results point to the fact that NBR’s medical utilization and its translation to clinical setting is emergent.^12^ For matrix reasoning however, only age affected DMN NBR. We believe that this result is related to the inherent difference in the difficulty of these two tasks as matrix reasoning is a harder task to perform, leading to saturation in the attenuation of NBR magnitude. As shown in Figure 4, the age-related decay in the magnitude of the NBR is approximately one-half SD for antonyms, whereas it is almost one SD for matrix reasoning, making further deterioration by Aβ deposition unlikely for this difficult task.

The pioneering work of Lustig et al described decreased DMN deactivation across aging and dementia of the Alzheimer type, however, their elderly cohort included participants with preclinical accumulation of Aβ (close to 30% of their older group), making it challenging to disentangle whether the observed effects were due to normal aging or early Aβ deposition. In addition, unlike our findings, they did not report any significant changes in Brodmann Area (BA 40) (Figures 1a/1b and Figures 2a/2b). Also, we did not identify any change in the direction of the BOLD response (from PBR to NBR or vice versa) for area BA 31as they had reported.

A similar paradigm was observed for the DAN PBR. The DAN PBR does not change significantly during antonyms, likely due to the simplicity of the task. In contrast, in the matrix reasoning task, age-related, but not Aβ-related, attenuation was observed in the PBR. This can be seen in the light of the DMN reaching a threshold resulting in an impairment of DAN activation for matrix reasoning. PBR from other networks showed similar results as they remained stable across aging and Aβ deposition (Supplementary Figures 1, 2, 3, and 5) or decreased (Supplementary Figure 6), except for limbic network during matrix reasoning task where HE showed significantly more PBR than HY (Supplementary Figure 4) warranting future investigation into these differences. While some studies have reported age-related^30,43,44^ and Aβ-related^45–47^ increases in the PBR, others have reported decreased activation with aging,^48–51^ or no significant association.^52^ Activation within the DAN is specifically a matter of dispute as there are reports of increased,^53,54^ decreased, ^55^ and preserved activation within the DAN.^56^ We focused on DAN PBR to clarify these discrepancies, showing a preserved PBR across healthy and pathological aging for antonyms but a reduced PBR across healthy aging for matrix reasoning. Reported age-related hyperactivation may reflect early vulnerability or inefficiency, rather than compensation, even driving Aβ deposition.^57^ PBR appears highly context-dependent and modulated by task difficulty, cognitive reserve, and subject-level factors.^58–60^

One crucial contributor to successful cognitive aging may be the preservation of a youth-like activation pattern as opposed to a reduced activation.^61^ While Chen et. al reported extra recruitment in successful aging, our elderly group outperformed the HY during antonyms, and their activation pattern (PBR) resembled the HY young group without additional recruitment. For the harder matrix reasoning task, the elderly group lost this young-like pattern, potentially impeding their performance. This supports the idea that maintaining an efficient youth-like pattern - not hyperactivation - underlies superior performance. Here, we showed an association between PBR and better performance despite the elderly group showing a youth-like pattern of activation during antonyms. The presence of such a pattern in our cognitively high functioning, socioeconomically advantaged elderly cohort may reflect a healthier brain phenotype and not a compensatory or pathological PBR pattern. Supporting this idea, prior studies have found that age-related brain atrophy can distort fMRI signals.^62^ Our findings suggest that PBR patterns might be subject-specific, varying with baseline health. Another explanation for why we did not observe a change in PBR across groups for antonyms might be because we report PBR as the multiplication of the BOLD expression by extent within DAN nodes that showed a significant PBR, however many studies which have reported hyperactivation discuss additional recruited areas showing only increased in overall PBR extent.^63^

### Robustness of FC networks across aging

Many fMRI studies have hypothesized that the deterioration in the FC networks may be the underlying mechanism that could explain the commonly reported age-related cognitive decline particularly in individuals free of Aβ burden especially in the higher-order networks such as the DMN^10,17,18,20,64,65^ and the DAN^17,18^ but also in the executive control network^17,18^ and the Salience network^17,18,66^ A mechanism for explaining the degradation of the FC networks can be age-related cortical atrophy as well as attenuating structural connectivity (SC) in an aging population ^67^ although FC and SC have shown to not always be directly related.^68^ This hypothesis is in contrast with more current findings that show lack of deterioration in FC or cognitive ability with age. In fact, some studies have shown the DMN FC to be very resilient and to maintain stability amid decreasing SC.^68^ One longitudinal study showed that DMN FC remains stable by age^69^, and another showed that if it does change, it only occurs at a regional or individual level but not on an average level.^70^ Similarly, another study showed stability of whole network DMN FC but only change within its individual subsystems.^71^ Other reports have shown different trends in FC depending on the adult age group (i.e middle vs. late age)^72^ or non-linear patterns across the lifespan.^73^ These contradictory findings along with other studies reporting preserved or even improved cognition with age^21,22^ weaken the argument that FC is directly linked to cognition at least at a whole network level.

Our results suggest that the FC of the whole DMN remains stable from young to older age with most regional pairs showing no significant change between HE and HY, consistent with more recent findings that show a lack of deterioration in FC with age. Three regional interhemispheric pairs within the temporal lobe showed lower DMN FC comparing HE versus HY. While these differences disappeared when head motion was constrained, these results should be investigated further to see how they may lose integrity with age and why they only occur for between hemisphere FC. Most interestingly, we have shown that not only the DMN FC, but other higher order networks also showed stability of within network FC suggesting that that age-related robustness of the FC networks may be a whole brain phenomenon and not restricted to a specific network. This is an important finding highlighting the resilience of the FC networks at a much broader level within the brain. One reason supporting the resilience of FC integrity can be attributed to a compensation hypothesis which suggests that the maintenance of the FC networks is desirable. A study from 2016 showed that FC within the cingulum bundle was preserved with age at the loss of SC and the two were not associated as hypothesized in the prior research.^68^ This study showed that FC can be maintained even with diminished underlying SC, and this may be necessary to maintain normal cognition. Possible explanations for the diversity in prior findings investigating age-related alteration in FC networks could be due to several factors including the lack of AD pathology measurements.^71^ Since AD pathology can be present in the brain decades before the onset of symptoms^5^, in-vivo measurement of Aβ burden may be necessary to disentangle whether the observed attenuation in the FC networks is due to normal aging or AD preclinical pathology. Since we obtained clinical amyloid readings for all participants included in the analysis, we believe our results are more reliable since we have distinct Aβ positivity readings for each elderly participant. Other considerations which may have contributed to conflicting findings in prior FC and aging studies can include interactions with tau pathology, differences in biomarker use, involuntary head motion during scanning, extensive brain atrophy, and cohort bias.

### Robustness of FC networks across preclinical AD

Whereas a clearer picture of the Aβ-FC relationship may exist in symptomatic AD, the relationship within pre-clinical AD has shown to be less definitive. For example, several past studies have reported significant findings related to the relationship between Aβ and FC within DMN regions. Some have shown decreasing FC^74–77^, some increasing,^78,79^ while some have reported both increasing or decreasing FC depending on the DMN region.^80^ A stage dependent effect was also reported within another study where hyperconnectivity was shown within the DMN during the earliest detectable stages of Aβ accumulation, followed by hypoconnectivity as Aβ burden increased further.^81^ Our results align most closely with the results from a study in 2016 which showed no significant associations between global Aβ burden and DMN FC in a middle-aged Hispanic population.^82^ Our study provides more evidence for the conclusions of Tahmi et. al for two reasons. The first is that we have included more ethnic groups in this study reducing potential cohort biases, and the second is that we compared amyloid Aβ to FC network changes across the seven major brain networks including the DMN and found similar results. We also replicated our findings within the DMN including different levels of framewise displacement to account more effectively for head motion and found no significant change in DMN FC with global amyloid. This evidence further supports the fact that Aβ and the FC networks on a whole network level may not be highly associated as previously thought.

### Association of BOLD response and FC with performance

In this study, we showed that FC and global Aβ uptake are not related to performance. However, aging, NBR, and PBR are associated with performance outcomes. This was true for both tasks, even though elderly participants performed better on antonyms but worse on matrix reasoning. Other papers have shown contradictory results on the nature of PBR and performance relationships. There are reports of better performance correlation with higher PBR from ventral lateral prefrontal cortex^83^, and DAN^58^ in studies using working memory tasks. These results are in line with our paper which showed that higher PBR within DAN is associated with better task performance and the fact that HE showed reduced PBR compared with the HY on matrix reasoning. These results suggest that PBR activation is task dependent and can differentiate between HY and HE when pushed harder using a more difficult task like matrix reasoning or like the two working memory tasks cited above that also used different loads.^84,85^ However, another study using executive task-switching experiments reported that higher activation predicted slower performance in elderly participants.^59^ Using a visual discrimination paradigm another study found that elderly with prefrontal overactivation performed poorly at the highest demand level.^58^ Although, these two studies also used different levels of difficulty, they are different from two other studies considering that they are executive tasks, again providing more evidence for the task-dependency of PBR.

Other studies have shown that increases in the magnitude of the DMN deactivation are usually directly related with better performance^49,51,52,54,55,57,61,63,64,85^ which is in line with what we show in our study for antonyms task. Interestingly, although elderly participants showed a reduced magnitude of NBR and a lower NBR magnitude itself, which is generally associated with worse performance, they still performed better on the antonyms task compared to healthy young. This might seem counterintuitive at first glance. However, prior studies have shown that performance on tasks like antonyms, which rely heavily on vocabulary knowledge, often sustain ^60^ or improve^86^ with age. Vocabulary tests are also considered proxies of cognitive reserve.^84,87,88^ Given that our participants were generally well educated and of higher socioeconomic status, with comparable overall cognitive abilities, we speculate that older adults’ better performance on the antonym task can be explained by the fact that this task relies on vocabulary knowledge and learning which accumulate throughout life. In contrast, matrix reasoning showed worse performance among elderly participants and no significant relationship with NBR, highlighting an inherent distinction. We propose that performance on crystalized memory (as tested by antonyms task) depend more on cognitive reserve and lifelong learning compared with matrix reasoning (a fluid reasoning task) that typically declines with age.

### Temporal ordering of the disruption of the FC and BOLD response

So far, we have discussed the stability of FC networks versus age- and Aβ-related attenuation of BOLD response in three group of participants (HY, HE, and CU). However, the within-subject alteration in the FC and BOLD response should also be investigated. Using a mixed-design repeated measure ANOVA we assessed the within subject differences between disruptions of NBR versus FC in the DMN as well as disruption of PBR and FC in DAN. This was only possible because our dataset included healthy young participant providing reference normative distribution for creating z-scores for alteration in FC and BOLD response. We showed that the DMN in each participant experiences age-related attenuation in the NBR, and this disruption worsens by presence of Aβ whereas FC of the same network remains stable in the same participant. By investigating the relationship between the subject-wise z-scores of the NBR and FC in the DMN and the bootstrap resampling of the posterior probabilities we showed that the probability of observing disruption in NBR when FC is already disrupted is significantly higher than probability of observing disruption in the FC when NBR is already disrupted. Altogether these results provide evidence that disruption in the NBR during aging and preclinical AD takes place before disruption in the FC of the DMN. There is one longitudinal study which reported that DMN FC remains stable over time in healthy aging, while deactivation reduces supporting our findings.^69^ However, this longitudinal study did not control for the presence of Aβ accumulation and its effect on FC, NBR, or PBR. Future studies with longitudinal imaging for NBR, FC, and Aβ is required to provide stronger evidence for the temporal ordering of the disruption of FC and BOLD responses in the brain networks.

### Limitations

Our dataset is cross-sectional and not optimal for investigating changes over time. Future longitudinal studies would be able to better discern a causal relationship between NBR and disease progression. These studies should investigate the relationship between baseline NBR and PBR and future Aβ accumulation. This helps to avoid possible challenge for studies such as ours that have included participants with higher education and socioeconomic class and comparable cognitive performance between young and old participants. Also considering that PBR from task positive networks shows no changes during antonyms and the fact that hyperactivation is observed in some studies, we believe DMN’s NBR is a more effective fMRI biomarker for exploring healthy and pathological conditions.^89^ Future studies should focus more on NBR as an important biomarker and investigate its relationship with tau and Aβ and the relationship between task difficulty and patterns of activation/deactivation to see if participants that are healthier at baseline and have an activation pattern like younger participants are less likely to accumulate Aβ in future.

## Conclusion

Our results provide evidence that brain functional networks go through an aging process which includes attenuation of task-evoked deactivation with potential loss of task-evoked activation depending on task difficulty. Pre-clinical accumulation of AD pathology contributed to further attenuation of task-evoked NBR which also was dependent on level of task difficulty. More importantly, we showed that the identified aging process for brain networks is the same for tasks improving or worsening with age suggesting a unified aging mechanism for the entire brain’s functional networks. Notably, the same networks FC did not show a significant alteration with age or preclinical AD pathology, suggesting a robust and resilient FC in the brain networks during normal aging and asymptomatic stage of AD. Similarly, the results of the within-subject analyses showed that each participant experiences a greater age-related alteration in the brain networks task-evoked BOLD response compared to FC and the difference between the two measurements revealed an interaction by the presence of AD pathology. Furthermore, we showed that within-subject alteration of the DMN task-evoked NBR most likely precedes alteration in underlying FC. Finally, we showed that task-evoked BOLD response is more closely associated with task performance than FC and global Aβ accumulation. Altogether, we conclude that the NBR from the DMN regions may be considered as an earlier and more reliable biomarker of brain functional networks disruption during aging and pre-clinical AD.

## Resource Availability

### Lead Contact

Requests for further information and resources should be directed to and will be fulfilled by the lead contact, Qolamreza R. Razlighi, PhD.

## Data and Code Availability

- Data reported in this paper will be shared by the lead contact upon request
- Any additional information required to reanalyze the data reported in this paper is available from the lead contact upon request.

## Data Availability

All data produced in the present study are available upon reasonable request to the authors

## Acknowledgments

We want to thank all the volunteers who selflessly participated in this project to help further the understanding of the brain during aging. Doing this project would not be possible without them.

## Author Contributions

Conceptualization: QRR, PC, BG

Methodology: QRR, PC, BG

Investigation: QRR, PC, BG, SHH, SO, JC, XHW, AM, SGP, GC

Visualization: PC, BG

Funding acquisition: QRR

Project administration: QRR, SO

Supervision: QRR

Writing – original draft: PC, BG

Writing – review & editing: QRR, PC, BG, SHH, SO, JC, XHW, AM, SGP, GC

## Supplementary Information

**Supplementary Table 1.** Mean functional connectivity for three different motion sub-samples within the DMN based on percentage of framewise displacement during rs-fMRI

**Supplementary Figure 1.** Task evoked positive BOLD response for Antonyms and functional connectivity changes across aging and pre-clinical AD within Limbic Network (LIM)

**Supplementary Figure 2.** Task evoked positive BOLD response for Antonyms and functional connectivity changes across aging and pre-clinical AD within Ventral Attention Network (VAN)

**Supplementary Figure 3.** Task evoked positive BOLD response for Antonyms and functional connectivity changes across aging and pre-clinical AD within Frontoparietal Network (FPN)

**Supplementary Figure 4.** Task evoked positive BOLD response for Matrix Reasoning and functional connectivity changes across aging and pre-clinical AD within Limbic Network (LIM)

**Supplementary Figure 5.** Task evoked positive BOLD response for Matrix Reasoning and functional connectivity changes across aging and pre-clinical AD within Ventral Attention Network (VAN)

**Supplementary Figure 6.** Task evoked positive BOLD response for Matrix Reasoning and functional connectivity changes across aging and pre-clinical AD within Frontoparietal Network (FPN)

**Supplementary Figure 7.** Results of statistical testing for significant differences in the mean of the inter-regional FC between the three groups controlling for motion (FD) (from left to right: HY-HE, HY-CU, and HE-CU)

## Material and Methods

### Study design

#### Participants screening and recruitment

Our cohort was primarily recruited within a 10-mile radius of the Citigroup Biomedical Imaging Facility (CBIC) at Weill Cornell Medicine (WCM) using random market mailing services between 2020 to 2024. All study procedures and data were collected at the CBIC facility within this timeframe. All participants in the study signed an informed consent form prior to enrollment and were compensated for their time spent on the study. The research experiments were performed in accordance with relevant guidelines and regulations approved by the WCM Institutional Review Board (IRB). Participants were screened by telephone. All enrolled participants were healthy, right-handed, and without MCI or dementia at the time of recruitment. This was confirmed during the visits by administering blind MoCA.

### Imaging procedures

#### Acquisition of MRI data

A research dedicated 3 Tesla Siemens Magnetom Prisma scanner with a 64 channel head-coil, and 80 mT/m gradient system was used in this study with a multiband T2* weighted echo-planar imaging (EPI) pulse sequence [TR/TE=1008/37 ms; flip angle=52°; FOV= 208 mm x 208 mm; matrix-size = 104 x 104; voxel-size = 2 x 2 x 2 mm; 72 axial slices; multiband factor = 6]. Each participant first underwent a scout localizer to determine the position and set the field of view and orientation, followed by high resolution T1-weighted magnetization-prepared rapid gradient-echo (MPRAGE) structural scan [TR/TE = 2400/3 ms; flip angle = 9°; FOV= 256’ 256 mm; matrix-size = 512 x 512; voxel-size = 0.5 x 0.5 x 0.5 mm; 320 axial slices] for localization and spatial normalization of the functional data in each participant. The duration of the tb-fMRI as well as rs-fMRI scan was 10 minutes. The phase encoding direction was alternated between consecutive rs-fMRI and tb-fMRI scans and used retrospectively for geometric distortion correction. In the case of any visual impairments, participants were instructed to use contact lenses, if possible, inside the scanner; otherwise, they were provided with MRI-safe visual aids.

#### ^18^F-Florbetaben Amyloid PET Imaging

Amyloid PET was performed in dynamic and 3D imaging mode on a Siemens Biograph mCT-S scanner. All participants’ vital signs were obtained before and after performing PET scanning with ^18^F-Florbetaben (FBB) PET tracer. Each participant’s preparation for the scans consisted of an intravenous (IV) catheterization, followed by the injection of 8.1 mCi ± 20% (300 MBq) of the tracer administered as a slow single IV bolus at 60 s or less (6 secs/mL max). Participants were scanned 90 minutes after the tracer injection. A low-dose CT scan for attenuation correction of the PET data was also acquired. Brain images for each of these PET scans were acquired in 4 × 5-minute frames over a 20-minute period.

#### PET Scan Visual Reading

Our trained and certified neuroradiologist on the study reviewed the images for amyloid deposition. The neuroradiologist was provided with de-identified images.

The images were then visually interpreted, and each participant’s scan was classified as positive or negative for amyloid at a global, whole-brain scale, and lobar regional level. Lobar characterization for each amyloid scan involved the evaluation of tracer uptake in the frontal, cingulate/precuneus, parietal, occipital, and temporal lobes. Visual readings were performed using *FSLeyes* by overlaying PET scans over the participant’s T1-weighted MPRAGE scan after rigid-body registration. The border of brain regions obtained by FreeSurfer were also available for the readers.

#### FreeSurfer Reconstruction of T1 scans

Each participant’s T_1_-weighted structural scan was reconstructed using FreeSurfer version 7 software package (http://surfer.nmr.mgh.harvard.edu/). This software segments cortical and subcortical areas based on the gyri and sulci morphology. The boundaries for grey/white matter and CSF were examined visually, slice by slice, by a single and blinded technician for each participant and if any visible discrepancy was detected, control points were added manually until the reconstruction results became satisfactory.^89^

#### Quantification Process for PET Data

An in-house developed pipeline for processing Amyloid PET scans was used; briefly by aligning dynamic PET frames (4 frames) to the first frame using rigid body registration. The structural T1 image in FreeSurfer was registered to the same participant’s static image using a rigid-body registration with normalized mutual information and 6 degrees of freedom. These transformed regional masks in static PET space were used to extract regional PET data. The standardized uptake value ratio (SUVR) was obtained by calculating the standardized uptake value (SUV) in FreeSurfer and Schaefer ROIs and then normalizing to the cerebellum gray matter. To attenuate the spill-in signal from nonspecific binding in white-matter, the uptake in the grey matter voxels located immediately adjacent to the white-matter volumes were discarded. This was done both in the cerebellum for computing the reference region uptake as well as in the cerebral cortex for obtaining cortical ROIs’ SUVR.

#### fMRI Pre-Processing

To address specific challenges associated with multiband-echo planar imaging (EPI) sequences, an in-house developed pre-processing pipeline was implemented to optimize data quality. Each participants’ fMRI native space was used in all pre-processing steps to prevent any inaccuracy in spatial normalization due to atrophy in the older and preclinical AD groups. The pre-processing sequence began with spatial realignment which was performed using FSL’s *mcflirt* to ensure accurate alignment of functional volumes. FSL’s *slicetimer* tool was then used for slice timing correction (temporal alignment). Spatial smoothing was applied using FSL’s *susan* tool with a 5 mm full width at half-maximum (FWHM). FSL’s ICA-based Automatic Removal of Motion Artifacts (AROMA) was employed to identify and remove the motion artifacts (the aggressive mode was used for rs-fMRI and non-aggressive mode for tb-fMRI data). The tb-fMRI scan and rs-fMRI scan acquired either before or after with opposite phase encoding direction were also fed to FSL’s topup tool to estimate the warping field. The FreeSurfer space brain mask was then inversely transformed to the participant’s fMRI space using the inverse of the rigid body transformation and the inverse of the geometric distortion correction warping field obtained with FSL’s *topup* tool. Asymmetric normalization tools (ANTS) were used to obtain a single warping field that transfers fMRI space to MNI template by performing a non-linear registration between the participant’s structural brain scans and MNI152 brain template. Global intensity normalization was performed after AROMA by making sure the global median of the fMRI data intensity (after removing outliers) was 10,000. Temporal filtering was performed next with high-pass band filter (f >0.01Hz). Finally, scrubbing was performed (FD > 0.5 mm, or global gray matter percent change signal RMSD>0.75).

### Cognitive Tasks

In this study, we have selected two different cognitive tasks from two cognitive domains (crystalized memory and fluid reasoning) to obtain the task-evoked networks of brain activation/deactivation. These two tasks were selected because one domain showed a significant age-related increase in performance, and the other domain showed an age-related decrease in performance. Both tasks were administered electronically with e-Prime 3 software (Psychology Software Tools, Pittsburgh, PA) using an event-related fMRI paradigm. Subjects were first trained outside of the scanner to learn and perform the tasks comfortably and accurately (∼30-60 minutes) prior to performing the actual fMRI session. The number of trials could not be balanced across tasks due to the differences in difficulty between tasks although the duration of the task versus the rest period was counterbalanced across tasks. All trial timings were jittered once for each task. The fMRI experiments were designed visually and presented within a square aligned to the horizontal meridian and were projected to a translucent screen located at the far end of the scanner. Each participant lied down supine inside the magnet bore, wearing noise isolating MR safe earbuds, and saw the screen using a mirror located on the head-coil. Participants responded to questions via a LUMItouch response system (Photon Control Company). The e-Prime software-controlled task administration collected response time (RT) and accuracy (ACC) metrics on a computer within the MRI control room. Task onset and MRI acquisition were electronically synchronized.

#### Antonyms

In this task, participants were asked to match a given word with its antonym or the word with the opposite meaning. An uppercase probe word was presented at the top of the screen inside a square, with four numbered choices presented below in lowercase and vertical order. Participants were instructed to respond quickly and accurately by pressing one of four buttons corresponding to the matching synonym. The task consisted of 40 trials with the following timing parameters: beginning fixation (BF)=7 sec; length of trial (LoT)=10 sec; inter-trial-interval (ITI)=uniform distribution (UD) [4.75, 14.75] second. Pre-training consisted of one set of 5 antonym questions.^90^

#### Matrix Reasoning

This task was adapted from the Raven matrix reasoning.^91^ During this task, participants identified a specific pattern from a figure series shown in a matrix with 3 rows and 3 columns. The matrix consisted of nine cells, with the bottom-right cell missing. Participants were presented with eight possible figures above and selected one of the five-figure choices that best completed the missing bottom right cell. For matrix reasoning, the timing parameters were as follows: BF=7s; TpR=15; LoT=35s; ITI=U [ [15.27,24.27] seconds. A pre-training session with each participant consisted of five practice problems. When asked, the correct answer was explained to the participant for each practice problem.

### Statistical Analyses

#### Functional Connectivity Analysis of fMRI

The same pre-processing pipeline for tb-fMRI was also used for FC data. The differences included the following: 1) no smoothing was applied, 2) the aggressive AROMA was used instead of non-aggressive, 3) a lower band-pass filter (0.01<f<0.1 Hz) was used instead of a high-pass filter. An inverse warp of the FreeSurfer segmentation into the fMRI native space was performed to extract white matter (WM) and ventricular signals along with motion parameters. These were regressed out from the time-series for all voxels. Schaefer atlas regional masks were then used to extract regional time-series by inversely warping them into the fMRI native space. Inter-regional FC was then computed by Pearson *C*orrelation Coefficient (PCC) between regional time series. Within network FC was computed by averaging all the pair-wise connections within each brain network from the Schaefer Atlas including the following: visual (VIS), somatomotor (SM), dorsal attention (DAN), ventral attention (VAN), limbic (LIM), frontoparietal (FPN), and default mode (DMN).

#### Subject-level and Group-Level fMRI Statistical Analysis

FSL’s *flim* tool was used to conduct subject-level statistical analyses. The general linear model (GLM) was applied to the pre-processed data to identify task-related neural activity. Design matrices included task timing box-cars convolved with the canonical double-gamma hemodynamic response function. Confound regressors included motion parameters and scrubbed volumes. The subject-wise PBR and NBR for each task were identified when defined contrasts of interest showed a significant difference from zero.^92^ First level statistical analysis results were held in the participant’s fMRI native space as well as warped to the MNI space for group-level analysis. Second level (group-level) analyses combined the MNI space statistical maps across participants using mixed-effects modeling (FLAME stages 1 and 2) Statistical thresholding was performed using cluster-based correction for multiple comparisons, with a cluster-forming threshold of Z>2.3 and a corrected cluster significance threshold of p<0.05. Z-scores for within network FC and within network NBR/PBR for older participants were created by normalizing to the mean and standard deviation of the healthy young group sample. Pairwise t-tests were performed to evaluate the difference in means between z-scores for within network FC and NBR/PBR. All pairwise comparisons for FC and NBR/PBR were adjusted for multiple comparisons using Bonferroni correction.

#### Cognitive Performance Analysis

Linear regression models were used to compare the performance on within-scanner crystalized memory (antonyms) and fluid reasoning (matrix reasoning) tasks. We used the ratio of accuracy to response time for each task as the outcome measure of performance. Four separate models were used per cognitive exam to identify associations in this outcome to four separate variables of interest. The main independent variables used for the four models were mean DMN NBR, mean DAN PBR, mean DMN FC, and Global Aβ SUVR, respectively. Each model was adjusted for both covariates (age, and gender).

#### Additional Statistical Analyses

All other statistical analyses were performed in R version 4.4.1 and python version 2.7.13. Clinical and demographic variables were analyzed by variable type. For all categorical variables in Table 1, chi-square tests were used to compare groupwise means and standard deviations. For all numerical variables, a Welch two sample t-test was performed to compare groupwise means and standard deviations. Linear regression was used to compare group differences in the mean of the FC for each network regions’ pair from Schaefer Atlas controlling for multiple comparison correction and individual participant FD in linear models. Pearson correlations (r) were performed to assess the degree of linear association of amyloid burden with network FC between groups. One-Way ANOVAs were performed to test for significant differences in mean NBR and PBR between the three groups and one-way ANCOVA when comparing mean FC since we adjusted for the individual framewise displacement (FD) of each participant. The *glht* (general linear hypothesis test) function in R, from the *multcomp* package, was used to assess pairwise comparisons in NBR and PBR between groups. For post-hoc tests, adjustment for multiple comparisons was performed using Bonferroni’s correction. Also, in post-hoc tests, we adjusted for age when comparing the two elderly groups with each other.

