## Supplemental Tables and Figures for "Age-enhancing cognitive ability shows similar attenuation in task evoked brain networks with aging and preclinical AD"

**Supplementary Figures and Tables**

**Supplementary Table 1.** Mean functional connectivity for three different motion sub-samples within the DMN based on percentage of framewise displacement during rs-fMRI

**Supplementary Figure 1.** Task evoked positive BOLD response for Antonyms and functional connectivity changes across aging and pre-clinical AD within Limbic Network (LIM)

**Supplementary Figure 2.** Task evoked positive BOLD response for Antonyms and functional connectivity changes across aging and pre-clinical AD within Ventral Attention Network (VAN)

**Supplementary Figure 3.** Task evoked positive BOLD response for Antonyms and functional connectivity changes across aging and pre-clinical AD within Frontoparietal Network (FPN)

**Supplementary Figure 4.** Task evoked positive BOLD response for Matrix Reasoning and functional connectivity changes across aging and pre-clinical AD within Limbic Network (LIM)

**Supplementary Figure 5.** Task evoked positive BOLD response for Matrix Reasoning and functional connectivity changes across aging and pre-clinical AD within Ventral Attention Network (VAN)

**Supplementary Figure 6.** Task evoked positive BOLD response for Matrix Reasoning and functional connectivity changes across aging and pre-clinical AD within Frontoparietal Network (FPN)

**Supplementary Figure 7.** Results of statistical testing for significant differences in the mean of the inter-regional FC between the three groups controlling for motion (FD) (from left to right: HY-HE, HY-CU, and HE-CU)

**Supplementary Figure 8.** Violin plots from replication analysis within the DMN for mean functional connectivity by motion for the following sub-samples (10%, 5%, and 1%) **Supplementary Table 1:** Within network mean DMN functional connectivity for each group by different levels of framewise displacement percentage

| Motion Sample | N=Total | N=Participants / Group  (HY, HE, CU) | HY | HE | CU |
| --- | --- | --- | --- | --- | --- |
| ≤ 10 % | 215 | N=55/125/35 | 0.399 | 0.392 | 0.378 |
| ≤ 5 % | 200 | N=52/116/32 | 0.387 | 0.389 | 0.374 |
| ≤ 1 % | 124 | N=35/67/22 | 0.350 | 0.373 | 0.365 |

**10 % Motion:** F (2, 213) =0.841 p=0.360, **5% Motion:** F (2,196) =0.266 **1% Motion:** F (1,122) =0.730 p=0.394

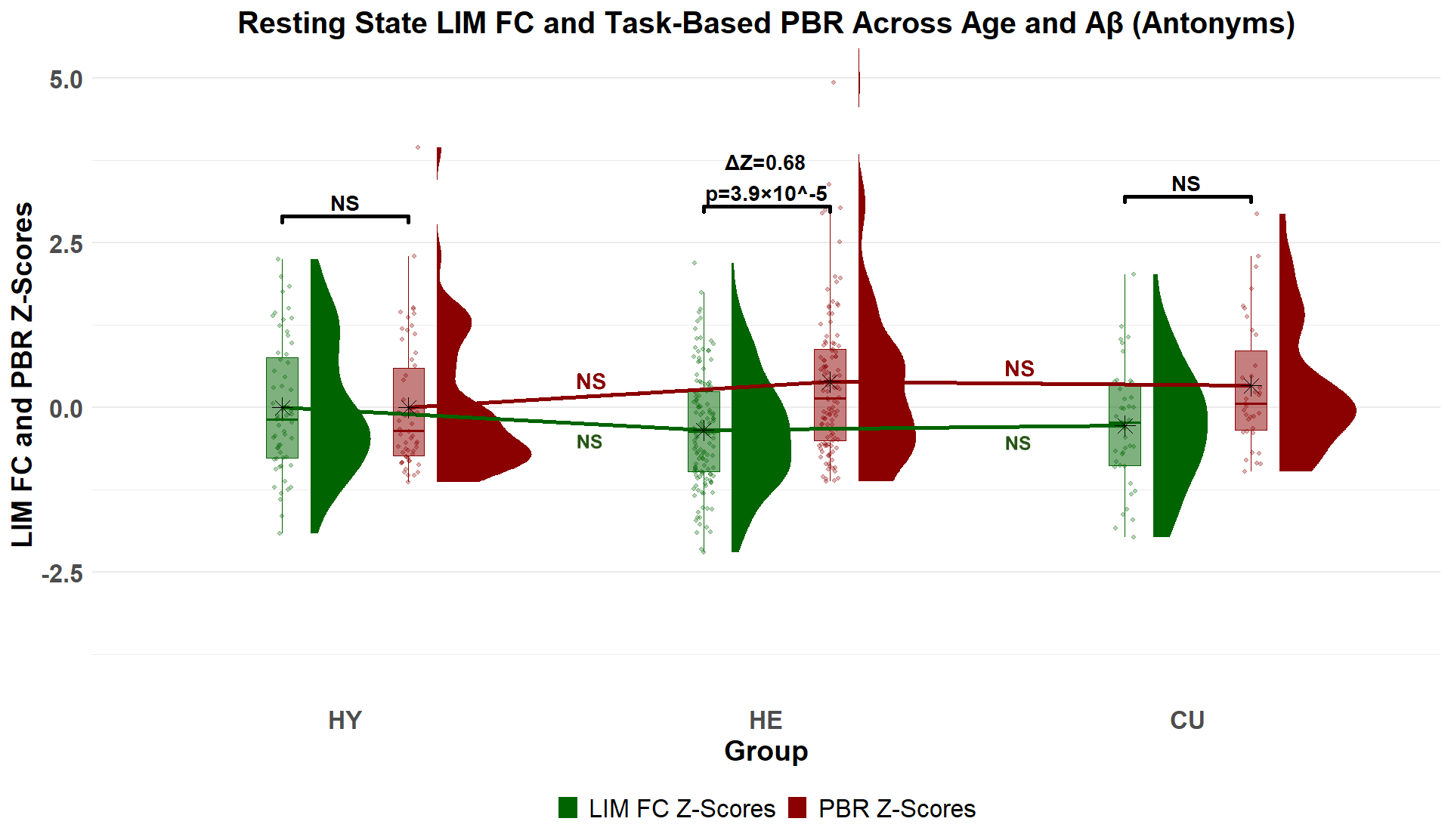

**Supplementary Figure 1. Limbic network’s age-related and Aβ-related alterations in the FC and task-evoked BOLD response during antonyms task,** Rain-cloud plots depict the distribution of the subject-wise positive BOLD response (in red) and FC (in green) z-scores for the HY, HE and CU groups. Within group differences stats are given in black.

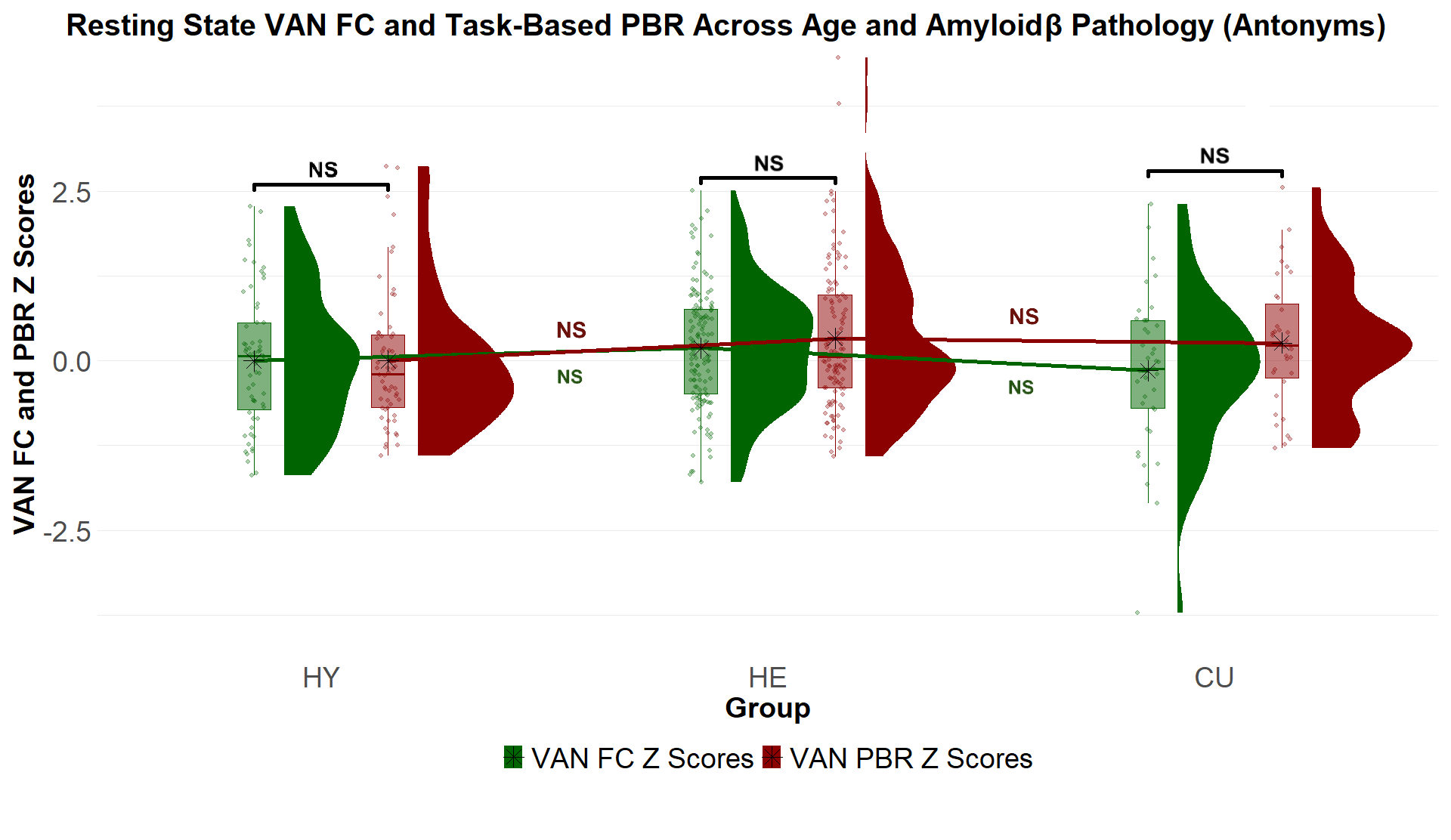

**Supplementary Figure 2. Ventral attention network’s age-related and Aβ-related alterations in the FC and task-evoked BOLD response during antonyms task;** Rain-cloud plots depict the distribution of the subject-wise positive BOLD response (in red) and FC (in green) z-scores for the HY, HE and CU groups. Within group differences stats are given in black.

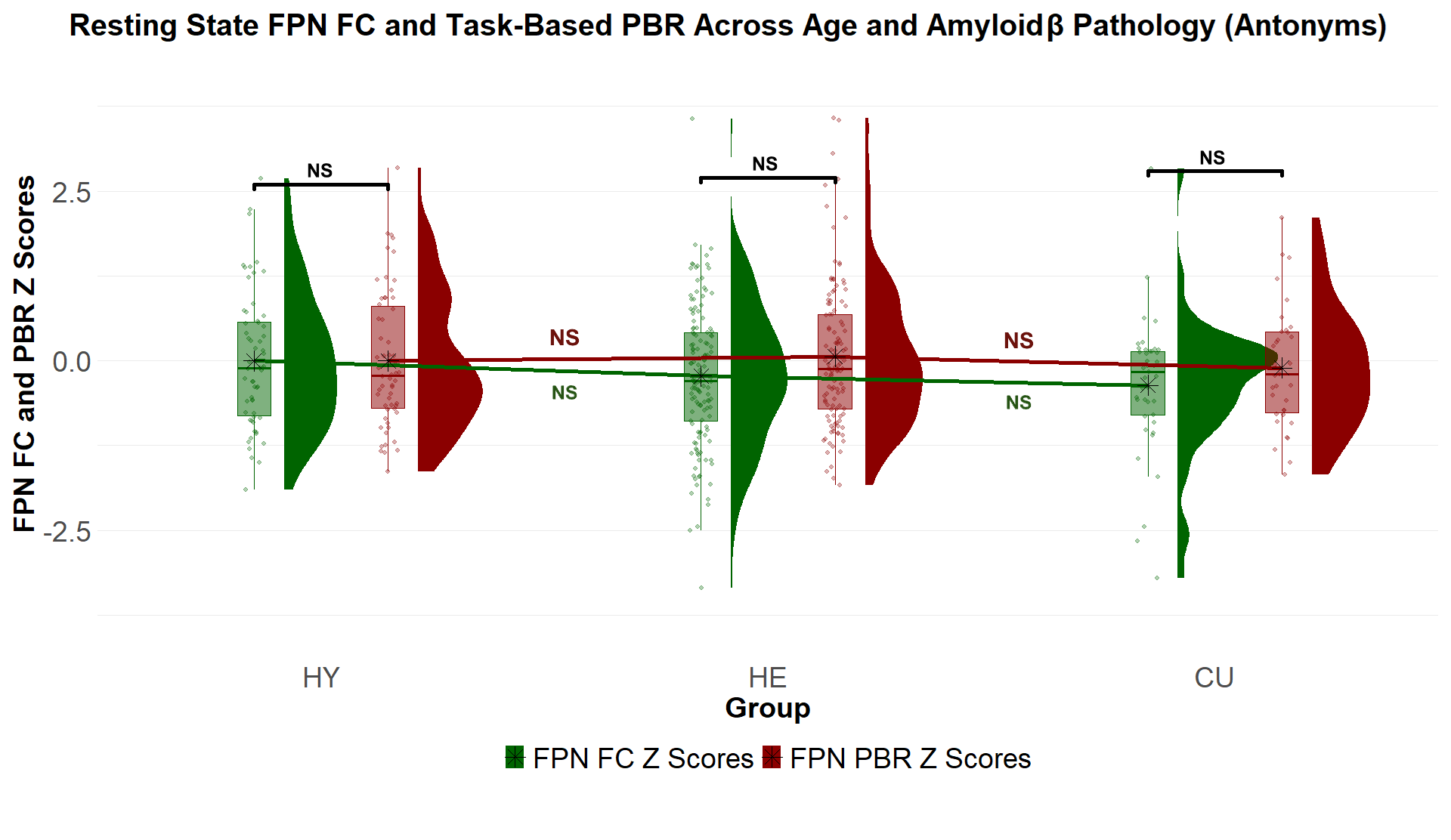

**Supplementary Figure 3. Frontoparietal network’s age-related and Aβ-related alterations in the FC and task-evoked BOLD response during antonyms task.** Rain-cloud plots depict the distribution of the subject-wise positive BOLD response (in red) and FC (in green) z-scores for the HY, HE and CU groups. Within group differences stats are given in black.

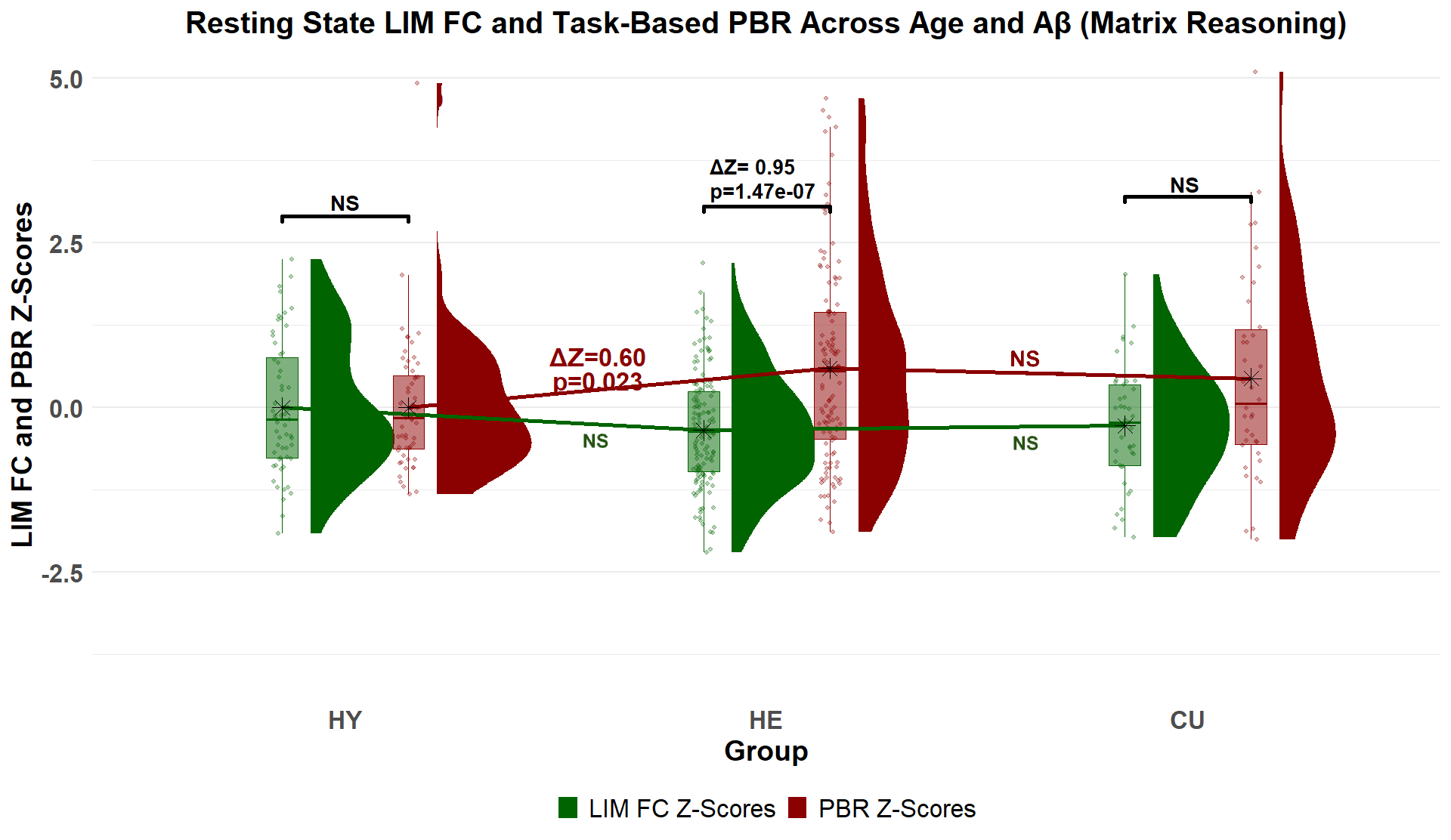

**Supplementary Figure 4. Limbic network’s age-related and Aβ-related alterations in the FC and task-evoked BOLD response during matrix reasoning task.** Rain-cloud plots depict the distribution of the subject-wise positive BOLD response (in red) and FC (in green) z-scores for the HY, HE and CU groups. Within group differences stats are given in black.

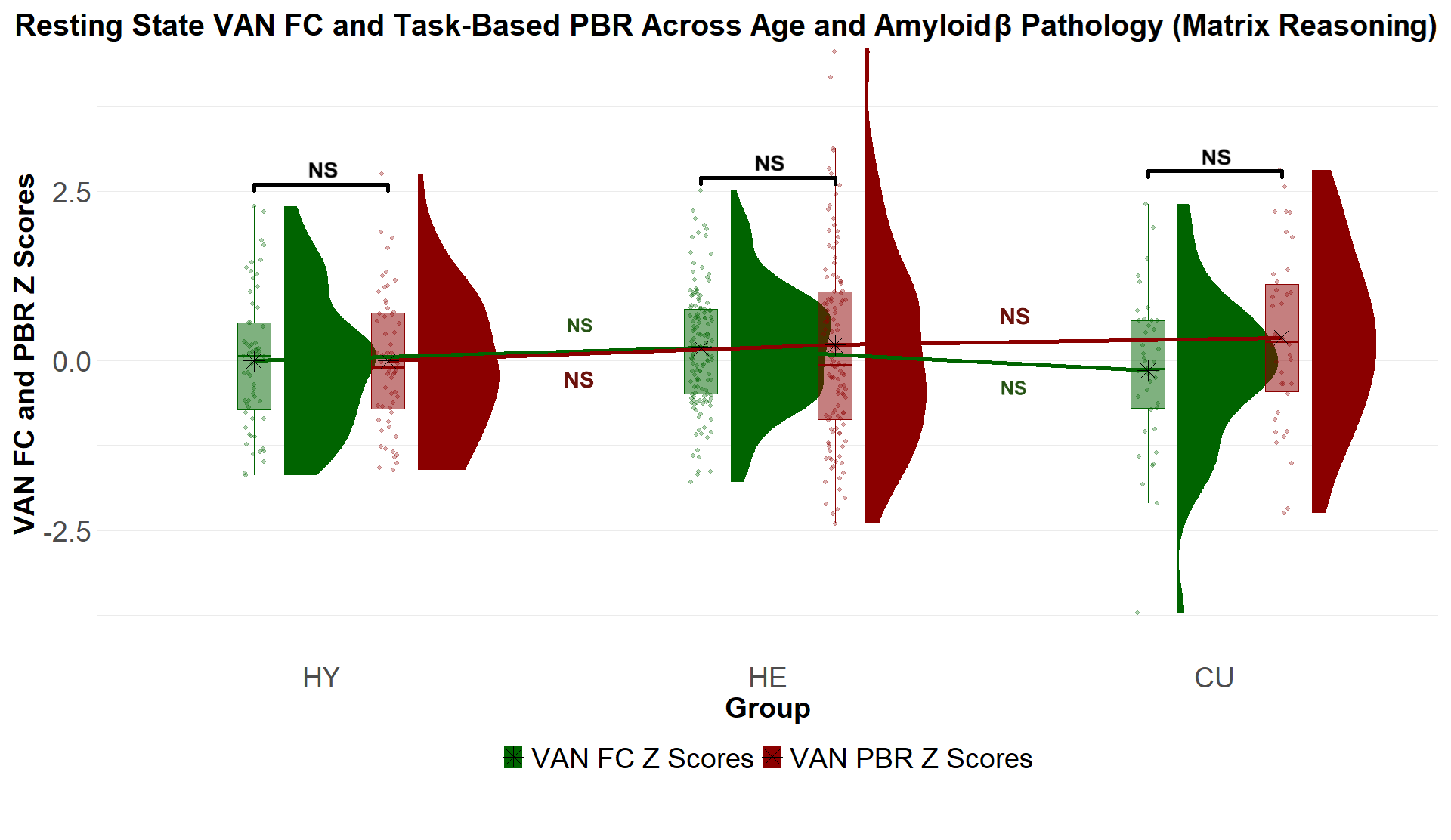

**Supplementary Figure 5. Ventral attention network age-related and Aβ-related alterations in the FC and task-evoked BOLD response during matrix reasoning task.** Rain-cloud plots depict the distribution of the subject-wise positive BOLD response (in red) and FC (in green) z-scores for the HY, HE and CU groups. Within group differences stats are given in black.

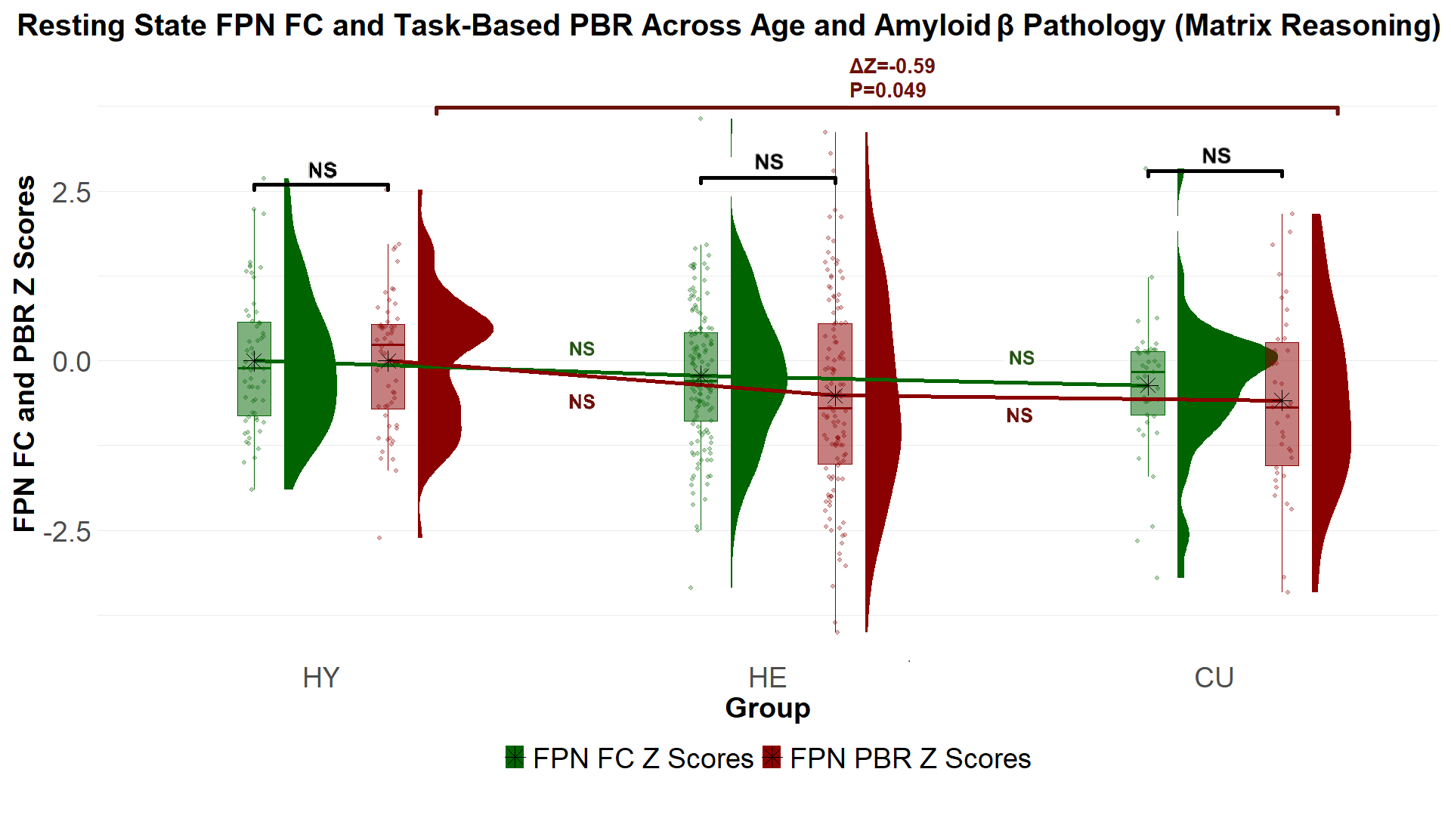
 **Supplementary Figure 6. Frontoparietal network age-related and Aβ-related alterations in the FC and task-evoked BOLD response during matrix reasoning task.** Rain-cloud plots depict the distribution of the subject-wise positive BOLD response (in red) and FC (in green) z-scores for the HY, HE and CU groups. Within group differences stats are given in black.

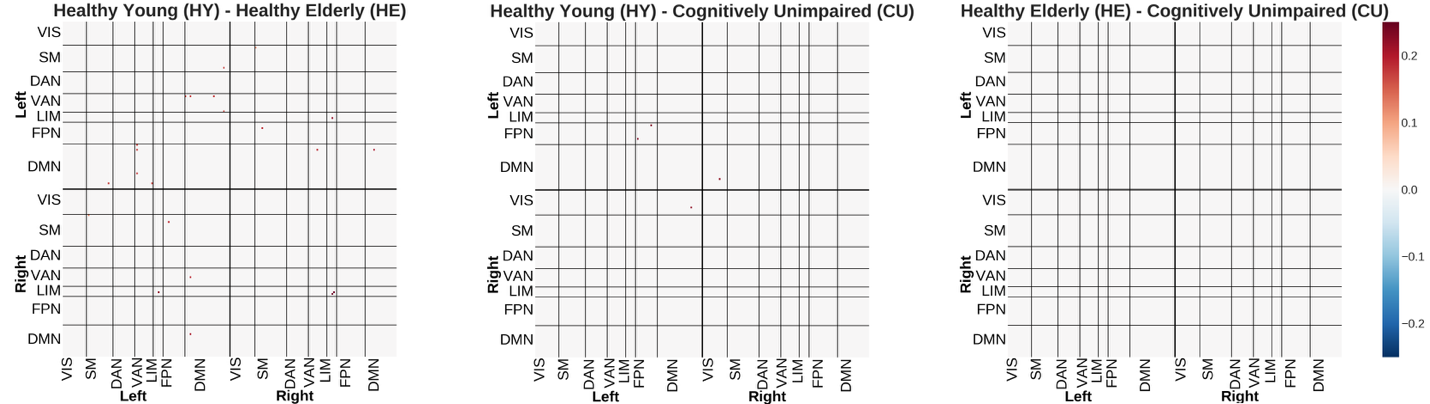

**Supplementary Figure 7. Results of statistical testing for significant differences in the mean of the inter-regional FC between the three groups controlling for motion (FD) (from left to right: HY-HE, HY-CU, and HE-CU)** The significant differences between mean FC that survived adjusting for motion and Bonferroni multiple comparison. No pairs survived the Bonferroni multiple comparisons for CU(Aβ+) vs HE(Aβ-) (right panel) and no pair in DMN survived for HY vs CU(Aβ+) (middle panel). However, a few pairs survived when comparing the FC of the HY vs HE(Aβ-) groups (left panel). Excluding participants with higher motions attenuated the significance level that did not survived multiple comparisons correction. The pair that survived Bonferroni multiple comparisons correction within the DMN was interhemispheric and within the temporal lobe (LH_Default_Temp_4 & RH_Default_Temp_3: p= 0.012), highlighting the possibility of the fMRI signal drop out in the temporal pole and inferior temporal regions.

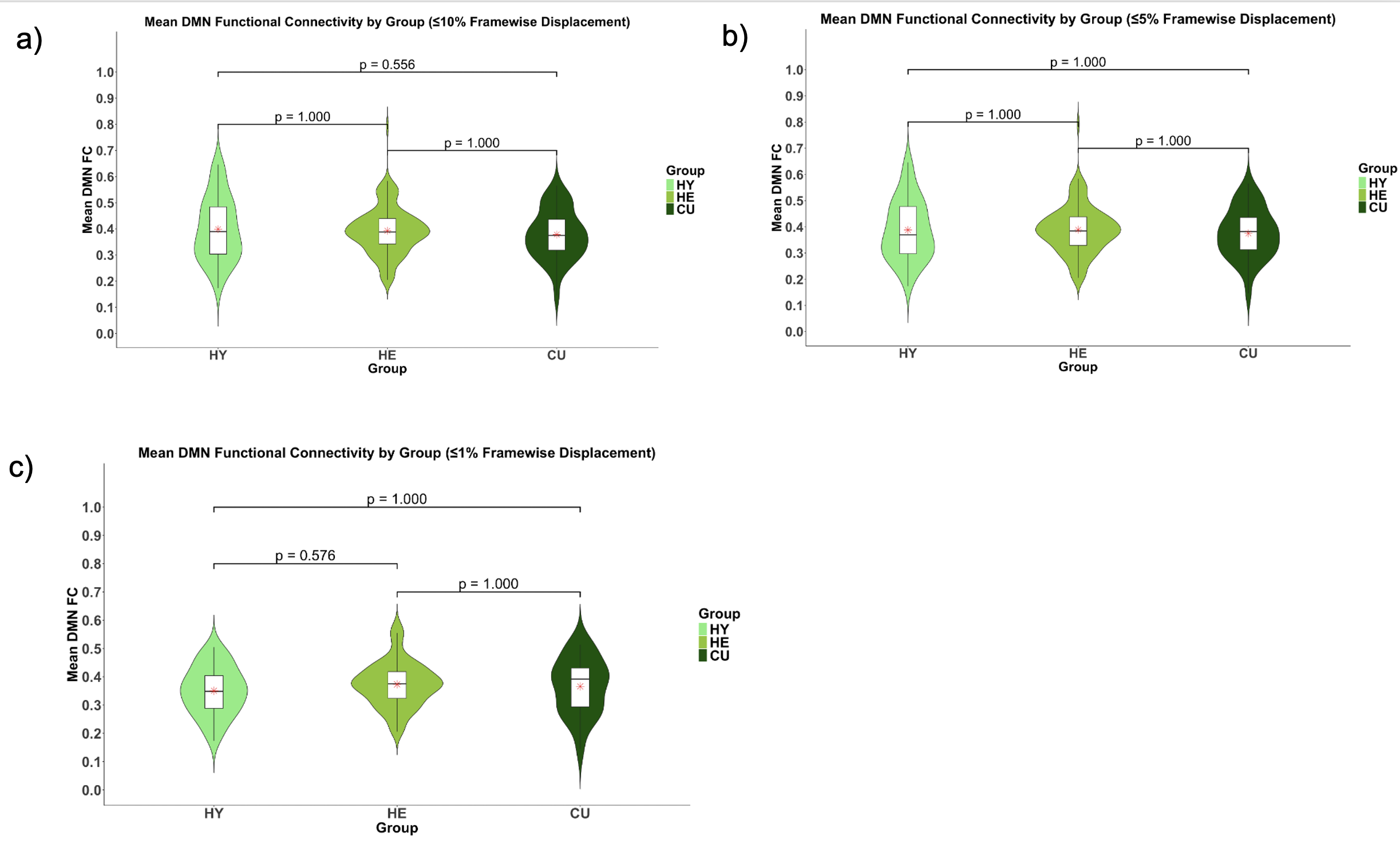

**Supplementary Figure 8.** **Violin plots from replication analysis within the DMN for mean functional connectivity by motion for the following sub-samples (10%, 5%, and 1%)** Distributions of subject-wise within network mean DMN FC for HY, HE, and CU groups for participants with different levels of motions categorized based on Framewise displacement (FD) in rs-fMRI a**)** FD ≤10%, b**)** FD ≤5%, and c) FD ≤1%
